# Understanding the pattern of cognitive decline in GBA1-related Parkinson’s Disease: a longitudinal multi-cohort study

**DOI:** 10.64898/2025.12.01.25341351

**Authors:** Merle Bode, Claire Pauly, Sonja Rós Jónsdóttir, Claudia Schulte, Sara Becker, Isabel Wurster, Benjamin Roeben, Stefanie Lerche, Werner Poewe, Rejko Krüger, Inga Liepelt-Scarfone, Kathrin Brockmann, the Parkinson’s Progression Markers Initiative* the NCER-PD consortium*

## Abstract

**Objective:** People with Parkinson’s disease (PD) who carry a pathogenic GBA1 variant (PD_GBA1_) are at higher risk of cognitive impairment than those without the variant (PD_GBA1_wildtype_). To date, little is known about the pattern and evolution of cognitive decline in PD_GBA1_. This multi-center study characterized the cognitive profile of PD_GBA_, focusing on the longitudinal trajectories and the group-specific onset times among cognitive functions, as well as their clinical relevance.

**Methods:** In this longitudinal multicohort-study (PPMI, ABC-PD, Luxembourg Parkinson’s Study), comprehensive neuropsychological assessments were standardized across 548 healthy controls (follow-up-years=2.84±4.33), 906 PD_GBA1_wildtype_ (follow-up-years=4.29±4.16), and 210 PD_GBA1_ (follow-up-years=4.09±3.35). We evaluated performance across age and disease duration using regression (generalized) linear mixed models within each cognitive domain. Time-to-first-event models, assessing risks of clinically relevant performance impairment (test-score *z*≤-1.5, MoCA<26), and an expanding-time-window approach identified the course of cognitive impairment. Additionally, correlations between cognitive functions were calculated.

**Results:** At baseline, PD_GBA1_ showed lower performance in attention (processing speed) and memory (verbal learning) than PD_GBA1_wildtype_, but more widespread probability of performance impairment. Over time, attention, visuoperception, memory, and semantic fluency performance declined more rapidly in PD_GBA1_ compared to PD_GBA1_wildtype_. Impairment in processing speed occurred earlier in the disease process of PD_GBA1_. Risks for clinically relevant cognitive impairment in PD_GBA1_ during the disease course were generally increased. Moderate-to-strong correlations between cognitive functions were observed within and across cognitive domains in PD_GBA1_ and PD_GBA1_wildtype_, particularly in attentional-executive functions.

**Interpretation:** PD_GBA1_ exhibits accelerated domain-generalized cognitive decline compared to PD_GBA1_wildtype_, with susceptibility of semantic fluency, attention, and memory.

**Summary for Social Media:**

1. <u>If you and/or a co-author has a X handle that you would like to be tagged, please enter it here. (format: @AUTHORSHANDLE)</u>. None
2. <u>What is the current knowledge on the topic? (one to two sentences)</u> Pathogenic coding variants in the Glucocerebrosidase (GBA1) gene in Parkinson’s Disease (PD-GBA1) are linked to more severe cognitive impairment. However, the domain-specific trajectories of cognitive decline are currently unknown.
3. <u>What question did this study address? (one to two sentences)</u> This study aimed to analyze the course of longitudinal cognitive profile in PD-GBA1 across disease duration and age in regard to cognitive domains and clinical relevance.
4. <u>What does this study add to our knowledge? (one to two sentences)</u> PD-GBA1 exhibits accelerated cognitive decline with susceptibility of semantic fluency, attention, and memory. Cognitive decline risk elevated 2-to-3 years post-diagnosis.
5. <u>How might this potentially impact on the practice of neurology? (one to two sentences)</u> The results can help to identify individuals at risk of cognitive decline and to better design clinical trials aiming at disease modification with cognition as endpoint.

## Introduction

Heterozygous variants in the gene encoding the enzyme β-glucocerebrosidase (GBA1)(1) are considered among the most clinically impactful genetic risk factors for Parkinson’s Disease (PD), with a prevalence of 14.3% in people with PD(2). Pathogenic coding GBA1 variants produce misfolded β-glucocerebrosidase enzymes, thereby impairing lysosomal-mediated alpha-synuclein degradation(3). People with PD who carry a pathogenic GBA1 variant (PD_GBA1_) present with an earlier and more severe onset of motor and non-motor symptoms than those who do not carry pathogenic coding GBA1 variants (PD_GBA1_wildtype_)(4,5).

Cognitive impairment, a troublesome and often-feared non-motor symptom in PD(6,7), is frequently experienced by people with PD_GBA1_(3). In general, PD_GBA1_ is linked to a higher risk of progressing to early dementia as compared to PD_GBA1_wildtype_(8,9). Knowledge of GBA1 variant status could motivate patients to make proactive health decisions(10) and could inform the development of disease-modifying clinical trial protocols(11). However, the understanding of the evolution and the temporal progression of the cognitive profile in PD_GBA1_ is limited.

Only two longitudinal studies have comprehensively examined the cognitive trajectories of PD_GBA1_(8,12). In such, a more severe decline in domains such as visuocognition, attention, executive function, and memory was reported(8,12). Although cross-sectional evidence shows some overlap with these findings, indicating that PD_GBA1_ perform worse in attention(13–15), executive functions(15,16), memory(17–19), and visuospatial skills(12,15,18,20), but not language(12–14,17,21), the timing and severity of the *individual cognitive functions* within these domains are reported inconsistently. This inconsistency raises the question of whether PD_GBA1_ represents a distinct clinical phenotype or an accelerated variant of PD_GBA1_wildtype_(12,22). Additionally, in conditions with widespread cognitive deficits such as PD_GBA1_, identifying primary sources of impairment can become challenging, since cognitive deficits in one function could affect performance on tests assessing other functions(23).

For clinical research and translation to clinical practice, it is important to determine whether statistical group differences between PD_GBA1_ and PD_GBA1_wildtype_ also represent clinically relevant impairments(24). Normative scores address this aim by quantifying the degree to which manifest cognitive performance deviates from what is expected for an individual’s education, sex, or age. In contrast, raw scores can be confounded by demographic factors, thus vulnerable to spurious group differences(25). To our knowledge, only two studies(17,20) have examined individual cognitive functioning using normative scores rather than domain composites(13,14,21), and none have systematically assessed whether cognitive performance crosses pathological thresholds.

Knowing *which* cognitive functions deteriorate *when* and at what associated *risk* in PD_GBA1_ relative to PD_GBA1_wildtype_ and healthy controls would enable the early identification of vulnerable patients, as well as the refinement and evaluation of disease-modifying trials(11). The goals of this study were to (1) characterize the decline of cognitive domains in PD_GBA1_ vs. PD_GBA1_wildtype_ vs. healthy controls over time, (2) classify the clinical relevance of cognitive decline, (3) evaluate the interrelations of cognitive domains among the study groups, and (4) explore the temporal course of cognitive domains impairment in PD_GBA1_ across disease duration.

## Methods

### Study design and participants

In this longitudinal multi-cohort study, we analyzed 1,664 participants (210 PD_GBA1_, 906 PD_GBA1_wildtype_, 548 healthy controls; see Fig. 1). Participants had been assessed within three cohorts: the international multicentric “The Parkinson’s Progression Markers Initiative (PPMI)” study(26), the monocentric “Amyloid-Beta in cerebrospinal spinal fluid as a risk factor for cognitive dysfunction in Parkinson’s Disease (ABC-PD)” study(27), and the monocentric “Luxembourg Parkinson’s Study” of the National Centre for Excellence in Research on Parkinson’s disease (NCER-PD)(28). The cohorts were harmonized based on overlapping cognitive tests. All studies adhered to the Declaration of Helsinki. All participants gave written informed consent, and all studies received ethical approval from ethics committees.

**Figure 1.**
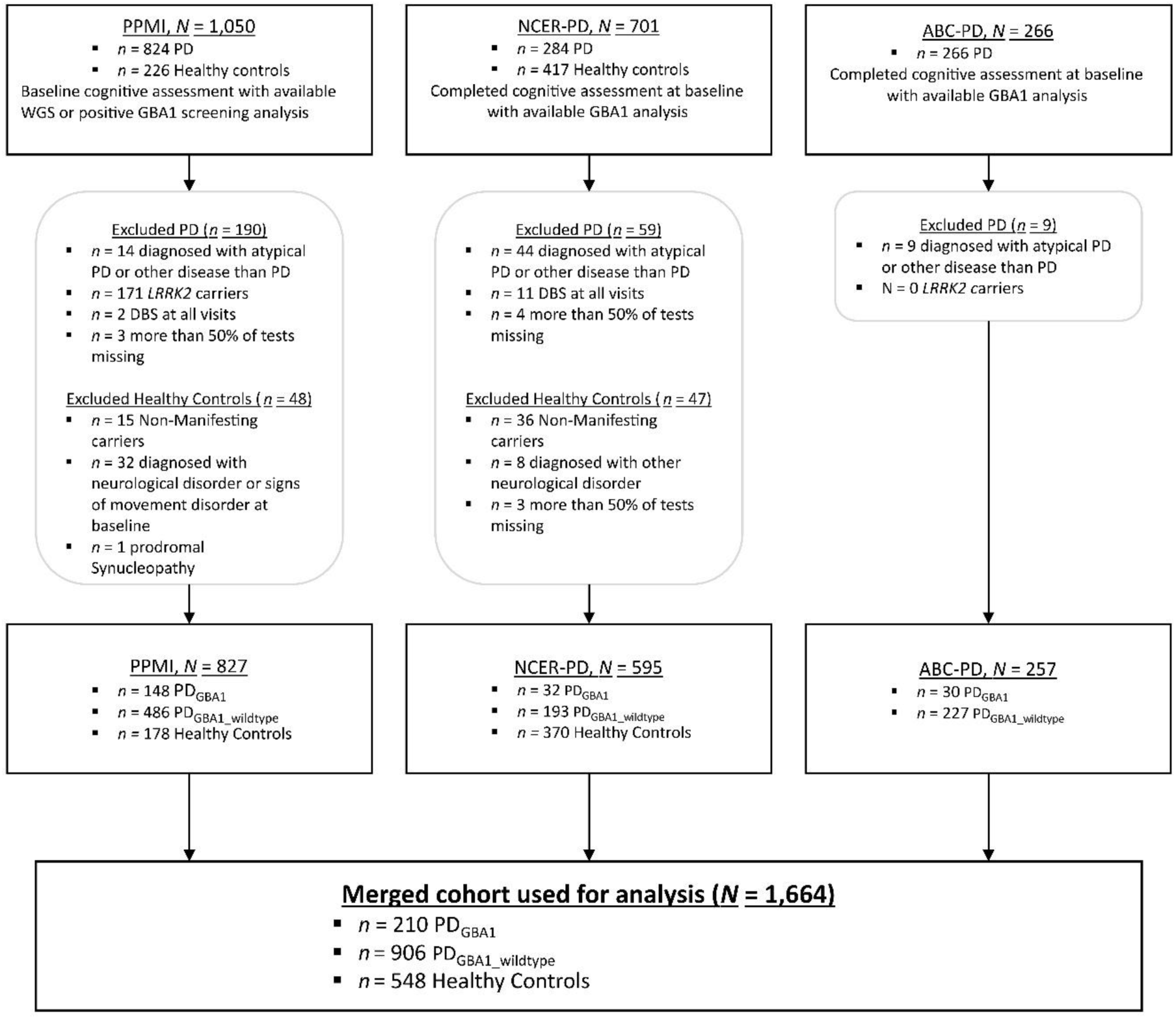
Flowchart of participant selection. *Note*. ABD-PD= “Amyloid-Beta in cerebrospinal spinal fluid as a risk factor for cognitive dysfunction in Parkinson’s Disease” study. DBS=Deep brain stimulation. GBA= Glucocerebrosidase gene. LuxPARK= “Luxembourg Parkinson’s Study at the University of Luxembourg”. PD=Parkinson’s Disease. PD_GBA_/_GBA1_wildtype_= people with PD carrying a/no pathogenic mutation in the Glucocerebrosidase gene. PPMI=Parkinson’s Disease Progression Marker Initiative. WGS=Whole genome sequencing.

The PPMI study is an ongoing international multicenter prospective longitudinal study of early de novo PD patients and healthy controls with annual visits. The study was launched in 2010 and is sponsored by the Michael J. Fox Foundation for Parkinson’s Research. Details of the study procedures have been described elsewhere(26), and study protocols can be found online at https://www.ppmi-info.org/study-design. Data was obtained from the PPMI database on the 22^nd^ of May 2025, and whole genome sequencing data was obtained on the 29^th^ of January 2025. The ABC-PD study is a closed monocentric prospective longitudinal cohort study of people with PD conducted at the University Hospital in Tübingen, Germany. The study included a baseline visit (between March 2014 and December 2017), a first follow-up visit (between July 2018 and December 2020), and a second follow-up visit (between March 2023 and January 2025). Details about the study procedures have been published elsewhere (27). The Luxembourg Parkinson’s Study is an ongoing nationwide monocentric prospective longitudinal cohort study on people with PD and healthy controls. PD participants are assessed annually, and healthy controls are followed up after 4 years. Details about the study procedures have been published elsewhere (28). Data was obtained on the12^th^ of February 2025.

Only participants with GBA1 variant analysis were included. The inclusion criteria for all participants were an absence of concomitant neurological diseases affecting cognition (e.g., stroke, traumatic brain injury, or encephalitis), and availability of at least 50% cognitive tests in each domain. *LRRK2* carriers were excluded from the analysis to avoid confounding due to protective effects of double-gene interactions(29). In the PD group, participants with a diagnosis or suspicion of atypical Parkinsonism or those who received deep brain stimulation were excluded. In the control group, GBA1 variant carriers (“non-manifesting”) and participants with signs of neurological disorders, including dementia or movement disorders, were excluded.

### Genetic Analysis

In PPMI, GBA1 variant status was defined based on whole genome sequencing data(26). To identify all GBA1 carriers, participants without available whole genome sequencing data but positive screening for the presence of GBA1 variants were included in the PD_GBA1_ group, but not those with negative screening, to avoid potential false negatives. Screening was obtained from genetic consensus data (https://ida.loni.usc.edu/), as well as GWAS, RNA-sequencing, Sanger sequencing of select GBA1 variants, and mutation screening data (“CLIA”). Consensus data included selected GBA1 variants: L483P (L444P), N409S (N370S), c.115+1G>A, and c.84dupG. In ABC-PD, genetic screening analysis for GBA1 variants was performed using the NeuroChip followed by Sanger sequencing for validation of the following GBA1 sequence variants: c.115+1G>A, R78C(R39C), T336S(T297S), E365K(E326K), R398*(R359X), T408M(T369M), N409S(N370S), and L483P(L444P)(27). In the Luxembourg Parkinson’s Study, screening was performed using a targeted PacBio DNA sequencing method and validated by Sanger sequencing(30).

### Assessments

Study assessments included demographics (age, sex, education, and disease duration), severity of motor symptoms and experiences of motor symptoms in daily life, assessed with part II and part III of the Movement Disorder Society (MDS) Unified Parkinson Disease Rating Scale (MDS-UPDRS)(31), and antiparkinsonian medication intake using levodopa equivalent daily dose (LEDD)(32). Depressive symptoms were measured with the Geriatric Depression Inventory(33) and the Beck Depression Inventory-I/II(34). Severity of depressive symptoms was classified in “normal”, “mild to moderate”, and “severe” based on cut-offs established by the respective manuals.

In comprehensive neuropsychological batteries, cognitive functions were assessed within the following domains: executive function (phonemic fluency, verbal fluency, set shifting), attention (working memory, processing speed), visuocognitive skills (visuoperception, visuoconstruction), memory (learning, delayed recall, retention, and discrimination), language, and global cognition (Montreal Cognitive Assessment; MoCA)(35). Cognitive functions were either represented by single tests or mean composites of multiple tests (see Table S1). In all tests, except for MoCA, raw performance was *z*-standardized based on published normative data, correcting for age, and whenever available, education and sex (Table S2). When published norms were unavailable, internal norms were generated using healthy controls as reference (Table S3). *Z*-scores were winsorized at *z*=-4.5.

Some tests became only available after the implementation of PPMI 2.0 and after 2018 in the Luxembourg Parkinson’s Study. Additionally, not all tests in the Luxembourg Parkinson’s Study were mandatory, with some being part of an optional assessment (Table S1). Specifically, this resulted in a lower number of available scores in cognitive functions such as visucoconstruction and set shifting. For a comprehensive cognitive analysis, these functions were included despite the smaller subsets in PD_GBA1_ (values missing set shifting: first visit=63%, last visit=20%, values missing visuoconstruction: first visit=79%, last visit=0%), PD_GBA1_wildtype_ (values missing set shifting: first visit=42%, last visit=13%, values missing visuoconstruction: first visit=71%, last visit=0%), and healthy controls (values missing set shifting: first visit=27%, last visit=0%, values missing visuoconstruction: first visit=84%, last visit=0%). For both cross-sectional and longitudinal analyses, statistical analyses were run using available data. Mean composites were calculated with available tests to minimize the number of missing values.

### Statistical analysis

Statistical analyses were performed using R (R Core Team, 2024). Alpha was set to α=.05. To maximize statistical power, both cross-sectional and longitudinal data was harmonized into a multi-panel structure, using chronological age and disease duration as common time scales. T-tests, Welch tests and χ2 tests were used, depending on data distribution, for between-group comparisons at the baseline visit and to identify potential confounding variables. Identified confounding variables were entered into stepwise forward elimination analyses. The selected confounding variables were included as covariates in all subsequent models. Missing values in confounding variables were handled using backward imputation and group-specific mean imputation within the harmonized cohort (Healthy controls, PD_GBA1_wildtype_, PD_GBA1_).

Cognitive functions were analyzed as *z*-scores or sum score for MoCA (“test performance”) or as binary indicators of test performance below established cut-offs (z≤-1.5) based on MDS criteria (24) based on MDS criteria, MoCA<26(, MoCA<26(35); “impaired performance”). Disease duration served as time scale in models, including direct comparisons between PD groups, allowing mapping of cognitive changes to disease duration. Chronological age served as time scale in models including comparisons of PD groups with healthy controls, thereby differentiating disease-related decline from normal aging. Between-group differences in the cognitive profile at the first study visit (=baseline) were assessed using linear or binomial regression models, including group main effects. Lower-order effects were only interpreted in absence of higher-order effects.

For Aim 1 (**to characterize cognitive trajectories**), linear mixed models (LMMs; “nlme 3.1”) were calculated to predict test performance of cognitive functions through time (chronological age, disease duration)*group interactions (PD groups vs. healthy controls, PD_GBA1_wildtype_ vs. PD_GBA1_). Intercepts for participants nested within cohorts were included as random effects. Intraclass correlations (“performance 0.13”)(36) measured homogeneity within study groups.

For Aim 2 (**to classify clinically relevant impairment**), binomial regressions were first computed to compare the occurrence of impaired performances among the three groups (PD groups vs. healthy controls, PD_GBA1_wildtype_ vs. PD_GBA1_). To account for variability in cognition, participants were classified as ever impaired in a cognitive outcome if impaired performance was detected at any available visit. The total number of visits was included as a covariate, as more frequent assessments could increase the chance of observing the outcome. Secondly, in line with MDS-Level-I criteria for PD mild cognitive impairment domain-level and global impairment were assessed. Binomial GLMMs (“glmmTMB 1.9”) were calculated in an analogous fashion to Aim 1, predicting the probability of impaired performance in each cognitive domain as well as the domains altogether (binary indicator; at least 1 z≤-1.5) and global cognition (MoCA<26).

For Aim 3 (**to evaluate the interrelations of cognitive functions**), correlational analyses were calculated between test performance on cognitive functions across the entire dataset.

For Aim 4 (**to explore the time course of cognitive impairment across disease duration**), the risks of impaired performance within each cognitive function were compared between PD groups (PD_GBA1_wildtype_ vs. PD_GBA1_) using time-to-first-event models (Cox regression; “survival 3.8”). The concordance index measured model fit. Subsequently, an expanding-time-window(37) was applied to the time-to-first-event models to identify when the risk of impaired performance first exceeded that of the reference group (“time of first deviation”, TFD): observation time windows were iteratively expanded across disease duration (1-year steps), with models re-estimated at each step until the risk within PD_GBA1_ first elevated from the risk of PD_GBA1_wildtype_ (HR>1.0, *p*<.05).

## 3. Results

### 3.1 Sample characterization

On average (Figure 2.a, Figure S1), PD_GBA1_ were followed for 4.09±3.35 years (visit range=1–13, Age range=34.30–88.60 years, Disease duration range=0–16.60 years), PD_GBA1_wildtype_ were followed 4.29±4.16 years (visit range=1–14, Age range=35.60–91.80 years, Disease duration range=0–29.40 years), and healthy controls were followed for 2.84±4.33 years (visit range=1–14, Age range=21.80–96.80 years).

**Figure 2.**
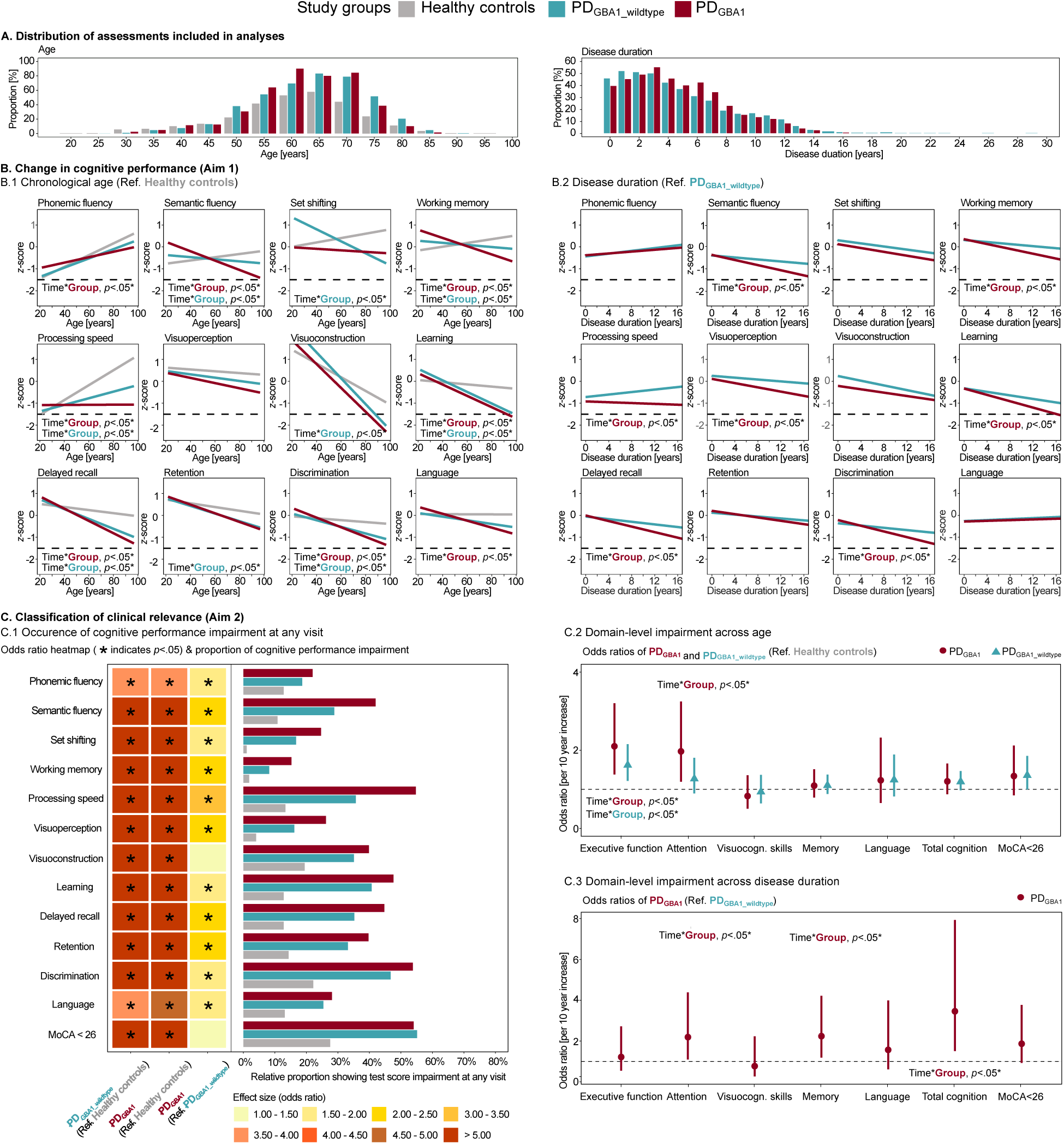
Overview of distribution of assessments and summaries of cognitive performance and in study groups (Aim 1, Aim 2) *Note.* Asterisks indicate *p*<.05. GBA= Glucocerebrosidase gene. PD=Parkinson’s Disease. PD_GBA/GBA1_wildtype_= people with PD carrying a/no pathogenic mutation in the Glucocerebrosidase gene. MoCA= Montreal Cognitive Assessment. *p*=p-value. Ref.=Reference group. Panel A.= shows the pooled age and disease duration distribution included in longitudinal analyses. Each bar represents the proportion of the baseline cohort (*n* = 548 healthy controls, *n* = 906 PD_GBA1_wildtype_, *n* = 210 PD_GBA_). Panel B.=Model predictions of linear mixed models assessing the trajectories of cognitive functions across age/disease duration per group (PD_GBA1_wildtype_ & PD_GBA1_ versus healthy controls, PD_GBA1_ versus PD_GBA1_wildtype_). Dashed lines represent the cut-off value indicating the clinical relevance of cognitive deficits (“test score impairment”). Panel C.1=The heatmap illustrates odds ratios from generalized linear models, indicating the likelihood of test score impairment at any available visit (“ever impaired”). The barchart illustrates the relative proportion of ever impaired participants within each study group. For example, 42% of participants within the PD_GBA1_ group showed a test score of *z*≤-1.5 in semantic fluency during at least one assessment in the study. Panels C.2/C.3=represent forest plots of odds ratios for domain-level impairment, as predicted by generalized linear mixed models over time (age, disease duration). A domain was classified as impaired if at least one test score of *z*≤-1.5 within the respective cognitive domain was observed, or if a MoCA of <26 was observed. Odds ratios are displayed per 10-year increase to simply interpretability of coefficients.

Demographic, clinical, and cognitive functions were compared between PD_GBA1_, PD_GBA1_wildtype_, and healthy controls (Table 1). Both PD_GBA1_ and PD_GBA1_wildtype_ were older (PD_GBA1_=62.98±9.78 years, *p*<.001; PD_GBA1_wildtype_=63.67±10.02 years, *p*<.001) than healthy controls (60.13±11.45 years) and more likely to report depressive symptoms (*p*<.001, respectively). Disease duration at baseline did not differ significantly between PD_GBA1_ (2.65±2.69 years) and PD_GBA1_wildtype_ (2.80±3.77 years). PD_GBA1_ were less likely to be male (*p*=.021), had higher education(*p*=.024), and received a higher LEDD (*p*=.002).

**Table 1.**
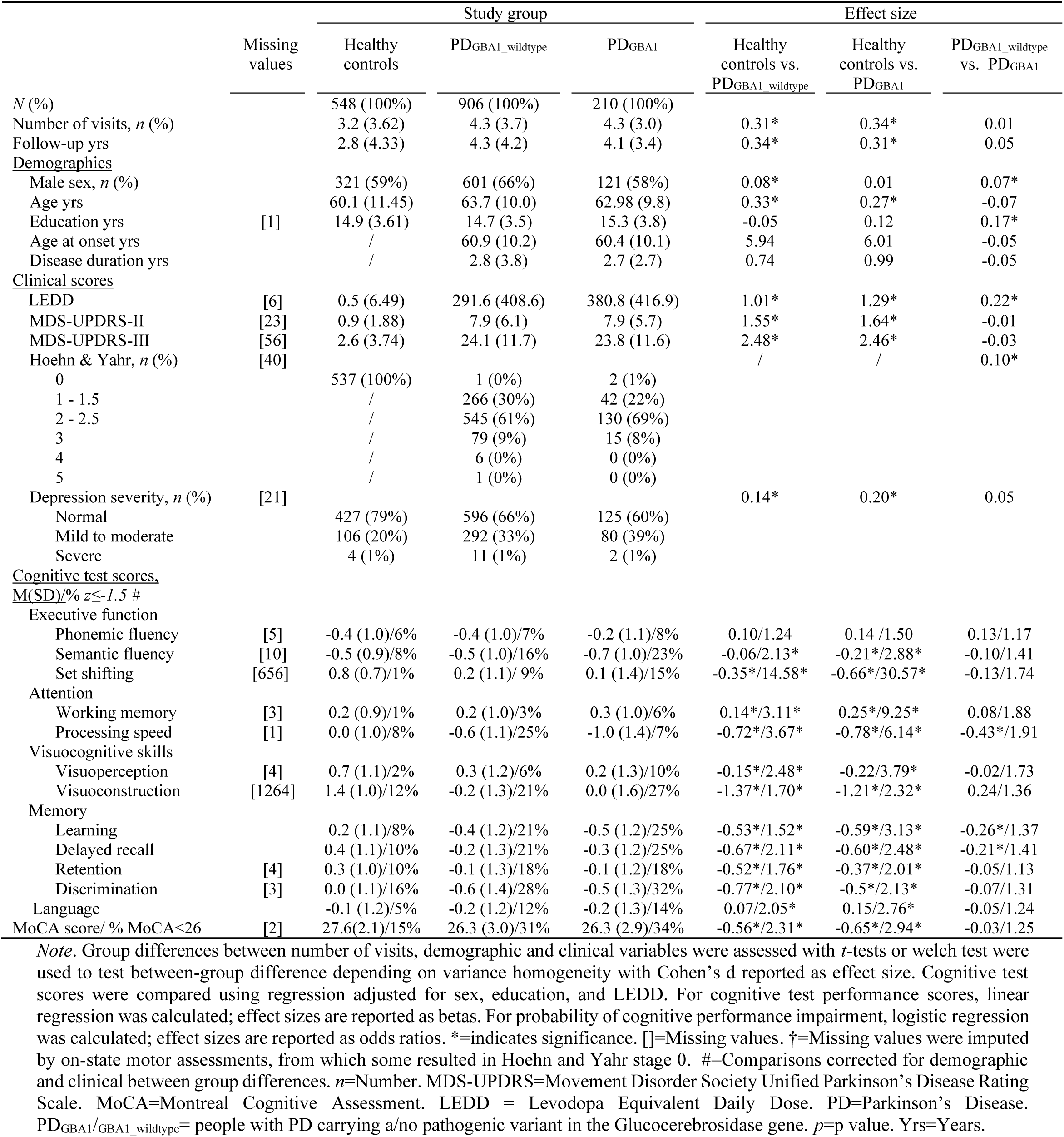
Baseline characterization of study sample.

|  | Missing values | Study group |  |  | Effect size |  |  |
| --- | --- | --- | --- | --- | --- | --- | --- |
|  |  | Healthy controls | PD<sub>GBA1</sub>_wildtype | PD<sub>GBA1</sub> | Healthy controls vs. PD<sub>GBA1</sub>_wildtype | Healthy controls vs. PD<sub>GBA1</sub> | PD<sub>GBA1</sub>_wildtype vs. PD<sub>GBA1</sub> |
| *N* (%) |  | 548 (100%) | 906 (100%) | 210 (100%) |  |  |  |
| Number of visits, *n* (%) |  | 3.2 (3.62) | 4.3 (3.7) | 4.3 (3.0) | 0.31* | 0.34* | 0.01 |
| Follow-up yrs |  | 2.8 (4.33) | 4.3 (4.2) | 4.1 (3.4) | 0.34* | 0.31* | 0.05 |
| <u>Demographics</u> |  |  |  |  |  |  |  |
| Male sex, *n* (%) |  | 321 (59%) | 601 (66%) | 121 (58%) | 0.08* | 0.01 | 0.07* |
| Age yrs |  | 60.1 (11.45) | 63.7 (10.0) | 62.98 (9.8) | 0.33* | 0.27* | -0.07 |
| Education yrs | [1] | 14.9 (3.61) | 14.7 (3.5) | 15.3 (3.8) | -0.05 | 0.12 | 0.17* |
| Age at onset yrs |  | / | 60.9 (10.2) | 60.4 (10.1) | 5.94 | 6.01 | -0.05 |
| Disease duration yrs |  | / | 2.8 (3.8) | 2.7 (2.7) | 0.74 | 0.99 | -0.05 |
| <u>Clinical scores</u> |  |  |  |  |  |  |  |
| LEDD | [6] | 0.5 (6.49) | 291.6 (408.6) | 380.8 (416.9) | 1.01* | 1.29* | 0.22* |
| MDS-UPDRS-II | [23] | 0.9 (1.88) | 7.9 (6.1) | 7.9 (5.7) | 1.55* | 1.64* | -0.01 |
| MDS-UPDRS-III | [56] | 2.6 (3.74) | 24.1 (11.7) | 23.8 (11.6) | 2.48* | 2.46* | -0.03 |
| Hoehn & Yahr, *n* (%) | [40] |  |  |  | / | / | 0.10* |
| 0 |  | 537 (100%) | 1 (0%) | 2 (1%) |  |  |  |
| 1 - 1.5 |  | / | 266 (30%) | 42 (22%) |  |  |  |
| 2 - 2.5 |  | / | 545 (61%) | 130 (69%) |  |  |  |
| 3 |  | / | 79 (9%) | 15 (8%) |  |  |  |
| 4 |  | / | 6 (0%) | 0 (0%) |  |  |  |
| 5 |  | / | 1 (0%) | 0 (0%) |  |  |  |
| Depression severity, *n* (%) | [21] |  |  |  | 0.14* | 0.20* | 0.05 |
| Normal |  | 427 (79%) | 596 (66%) | 125 (60%) |  |  |  |
| Mild to moderate |  | 106 (20%) | 292 (33%) | 80 (39%) |  |  |  |
| Severe |  | 4 (1%) | 11 (1%) | 2 (1%) |  |  |  |
| <u>Cognitive test scores, M(SD)/% $z \leq -1.5$ #</u> |  |  |  |  |  |  |  |
| <u>Executive function</u> |  |  |  |  |  |  |  |
| Phonemic fluency | [5] | -0.4 (1.0)/6% | -0.4 (1.0)/7% | -0.2 (1.1)/8% | 0.10/1.24 | 0.14 /1.50 | 0.13/1.17 |
| Semantic fluency | [10] | -0.5 (0.9)/8% | -0.5 (1.0)/16% | -0.7 (1.0)/23% | -0.06/2.13* | -0.21*/2.88* | -0.10/1.41 |
| Set shifting | [656] | 0.8 (0.7)/1% | 0.2 (1.1)/ 9% | 0.1 (1.4)/15% | -0.35*/14.58* | -0.66*/30.57* | -0.13/1.74 |
| <u>Attention</u> |  |  |  |  |  |  |  |
| Working memory | [3] | 0.2 (0.9)/1% | 0.2 (1.0)/3% | 0.3 (1.0)/6% | 0.14*/3.11* | 0.25*/9.25* | 0.08/1.88 |
| Processing speed | [1] | 0.0 (1.0)/8% | -0.6 (1.1)/25% | -1.0 (1.4)/7% | -0.72*/3.67* | -0.78*/6.14* | -0.43*/1.91 |
| <u>Visuocognitive skills</u> |  |  |  |  |  |  |  |
| Visuoperception | [4] | 0.7 (1.1)/2% | 0.3 (1.2)/6% | 0.2 (1.3)/10% | -0.15*/2.48* | -0.22/3.79* | -0.02/1.73 |
| Visuoconstruction | [1264] | 1.4 (1.0)/12% | -0.2 (1.3)/21% | 0.0 (1.6)/27% | -1.37*/1.70* | -1.21*/2.32* | 0.24/1.36 |
| <u>Memory</u> |  |  |  |  |  |  |  |
| Learning |  | 0.2 (1.1)/8% | -0.4 (1.2)/21% | -0.5 (1.2)/25% | -0.53*/1.52* | -0.59*/3.13* | -0.26*/1.37 |
| Delayed recall |  | 0.4 (1.1)/10% | -0.2 (1.3)/21% | -0.3 (1.2)/25% | -0.67*/2.11* | -0.60*/2.48* | -0.21*/1.41 |
| Retention | [4] | 0.3 (1.0)/10% | -0.1 (1.3)/18% | -0.1 (1.2)/18% | -0.52*/1.76* | -0.37*/2.01* | -0.05/1.13 |
| Discrimination | [3] | 0.0 (1.1)/16% | -0.6 (1.4)/28% | -0.5 (1.3)/32% | -0.77*/2.10* | -0.5*/2.13* | -0.07/1.31 |
| Language |  | -0.1 (1.2)/5% | -0.2 (1.2)/12% | -0.2 (1.3)/14% | 0.07/2.05* | 0.15/2.76* | -0.05/1.24 |
| MoCA score/ % MoCA<26 | [2] | 27.6(2.1)/15% | 26.3 (3.0)/31% | 26.3 (2.9)/34% | -0.56*/2.31* | -0.65*/2.94* | -0.03/1.25 |

*Note.* Group differences between number of visits, demographic and clinical variables were assessed with *t*-tests or welch test were used to test between-group difference depending on variance homogeneity with Cohen's *d* reported as effect size. Cognitive test scores were compared using regression adjusted for sex, education, and LEDD. For cognitive test performance scores, linear regression was calculated; effect sizes are reported as betas. For probability of cognitive performance impairment, logistic regression was calculated; effect sizes are reported as odds ratios. \*=indicates significance. []=Missing values. †=Missing values were imputed by on-state motor assessments, from which some resulted in Hoehn and Yahr stage 0. #=Comparisons corrected for demographic and clinical between group differences. *n*=Number. MDS-UPDRS=Movement Disorder Society Unified Parkinson's Disease Rating Scale. MoCA=Montreal Cognitive Assessment. LEDD = Levodopa Equivalent Daily Dose. PD=Parkinson's Disease. PD<sub>GBA1</sub>/GBA1\_wildtype= people with PD carrying a/no pathogenic variant in the Glucocerebrosidase gene. *p*=*p* value. Yrs=Years.

Compared with healthy controls (Table 1), PD_GBA1_ scored significantly worse across nearly all cognitive functions, except phonemic fluency, visuoperception, and language. PD_GBA1_wildtype_ also exhibited lower cognitive performance than healthy controls across various functions, though their performance in phonemic and semantic fluency, as well as language, was similar to that of healthy controls. Compared to PD_GBA1_wildtype_, PD_GBA1_ demonstrated slower processing speed, decreased memory, learning capacity, and delayed recall at baseline.

Impaired performance (Table 1) in the MoCA was observed in 34% of PD_GBA1_, 31% in PD_GBA1_wildtype_, and 15% in healthy controls. In the other cognitive functions, impaired performance ranged from 6% (working memory) to 37% (processing speed) in the PD_GBA1_ group, from 3% (working memory) and 28% (discrimination) in the PD_GBA1_wildtype_ group, and from 1% (set shifting, working memory) to 12% (visuoconstruction) in the healthy control group. Binomial regression indicated that the probability of impaired performance for both PD_GBA1_ and PD_GBA1_wildtype_ was increased across all functions compared to healthy controls, except for phonemic fluency. Compared to PD_GBA1_wildtype_, PD_GBA1_ showed a significantly higher probability of impaired performance in nearly all functions, except for phonemic fluency, retention, and language.

### 3.2 Aim 1: Change in cognitive test performance over time (test performances)

Longitudinal trajectories in test performance were compared between study groups using LMMs (Table 2; detailed results reported in Table S4-S5). Using age as time scale to differentiate disease effects from age-related changes (Figure 2.b.1), both PD_GBA1_ and PD_GBA1_Wildtype_ showed a steeper decline than healthy controls in MoCA, executive function (phonemic fluency, semantic fluency), attention (working memory, processing speed), and memory performance (learning, recall, discrimination). Additionally, PD_GBA1_ showed a steeper decline in language performance compared with healthy controls. Only PD_GBA1_wildtype_ showed steeper decline than healthy controls in set shifting, visuoconstruction, and retention. Intraclass correlations (Table S6) indicated fair to good homogeneity (42%–74%) across all study groups and functions, except for MoCA, for which borderline poor homogeneity was observed in the PD_GBA1_ group (39%). Using disease duration as time scale to map cognitive changes to disease progression (Figure 2.b.2), PD_GBA1_ showed significantly steeper decline than PD_GBA1_wildtype_ in executive functions (semantic fluency), attention (working memory and processing speed), visuocognitive skills (visuoperception), and memory (learning, delayed recall, discrimination). Intraclass correlations indicated fair to good homogeneity (41%–72%) across all study groups and functions.

**Table 2.** Results of linear mixed effects models (LMMs) predicting cognitive impaired performance (*z≤* -1.5, MoCA<26) by time*study group interactions across chronological age and disease duration.

|  | Age |  |  |  | Disease duration |  |
| --- | --- | --- | --- | --- | --- | --- |
|  | PD<sub>GBA1</sub> wildtype vs. Healthy controls |  | PD<sub>GBA1</sub> vs. Healthy controls |  | PD<sub>GBA1</sub> vs. PD<sub>GBA1</sub> wildtype |  |
|  | Beta [95% CI] | *p* | Beta [95% CI] | *p* | Beta [95% CI] | *p* |
| Executive function |  |  |  |  |  |  |
| Phonemic fluency | -0.01 [-0.01, 0.00] | .065 | -0.02 [-0.03, 0.00] | .010 * | -0.01 [-0.03, 0.01] | .274 |
| Semantic fluency | -0.01 [-0.02, 0.00] | .002 * | -0.03 [-0.04, -0.02] | <.001 * | -0.04 [-0.06, -0.02] | <.001 * |
| Set shifting | -0.04 [-0.05, -0.03] | <.001 * | -0.01 [-0.03, 0.00] | .148 | -0.01 [-0.05, 0.03] | .672 |
| Attention |  |  |  |  |  |  |
| Working memory | -0.01 [-0.02, -0.01] | <.001 * | -0.03 [-0.04, -0.02] | <.001 * | -0.03 [-0.05, -0.02] | <.001 * |
| Processing speed | -0.02 [-0.03, -0.01] | <.001 * | -0.04 [-0.05, -0.02] | <.001 * | -0.04 [-0.06, -0.02] | <.001 * |
| Visuocognitive skills |  |  |  |  |  |  |
| Visuoperception | 0.00 [-0.01, 0.01] | .414 | -0.01 [-0.02, 0.01] | .254 | -0.03 [-0.05, 0.00] | .025 * |
| Visuoconstruction | -0.03 [-0.04, -0.01] | .005 * | -0.02 [-0.05, 0.01] | .230 | 0.02 [-0.05, 0.08] | .633 |
| Memory |  |  |  |  |  |  |
| Learning | -0.02 [-0.03, -0.01] | <.001 * | -0.02 [-0.04, -0.01] | .001 * | -0.03 [-0.06, -0.01] | .005 * |
| Delayed recall | -0.02 [-0.02, -0.01] | <.001 * | -0.02 [-0.04, -0.01] | .001 * | -0.03 [-0.06, -0.01] | .005 * |
| Retention | -0.01 [-0.02, 0.00] | .042 * | -0.01 [-0.02, 0.00] | .095 | -0.02 [-0.04, 0.01] | .243 |
| Discrimination | -0.01 [-0.02, 0.00] | .016 * | -0.02 [-0.03, 0.00] | .011 * | -0.04 [-0.07, -0.01] | .008 * |
| Language | -0.01 [-0.02, 0.00] | .095 | -0.02 [-0.03, 0.00] | .029 * | 0.00 [-0.03, 0.02] | .788 |
| MoCA score | -0.06 [-0.08, -0.04] | <.001 * | -0.08 [-0.11, -0.04] | <.001 * | -0.04 [-0.10, 0.01] | .114 |

*Note.* Interaction coefficients represent change per year of age/disease duration in PD<sub>GBA1</sub>/PD<sub>GBA1</sub> in respect to the reference group. For LMMs across chronological age, sex, education, LEDD, and depression severity were used as covariates. For LMMs across disease duration, sex, education, and LEDD were used as covariates. Asterisks indicate significance. 95%-CI= 95% Confidence Interval. MoCA=Montreal Cognitive Assessment. PD=Parkinson's Disease. PD<sub>GBA1</sub>/G<sub>BA1</sub>\_wildtype= people with PD carrying a/no pathogenic variant in the Glucocerebrosidase gene. *p*=*p* value.

**Table 3.**
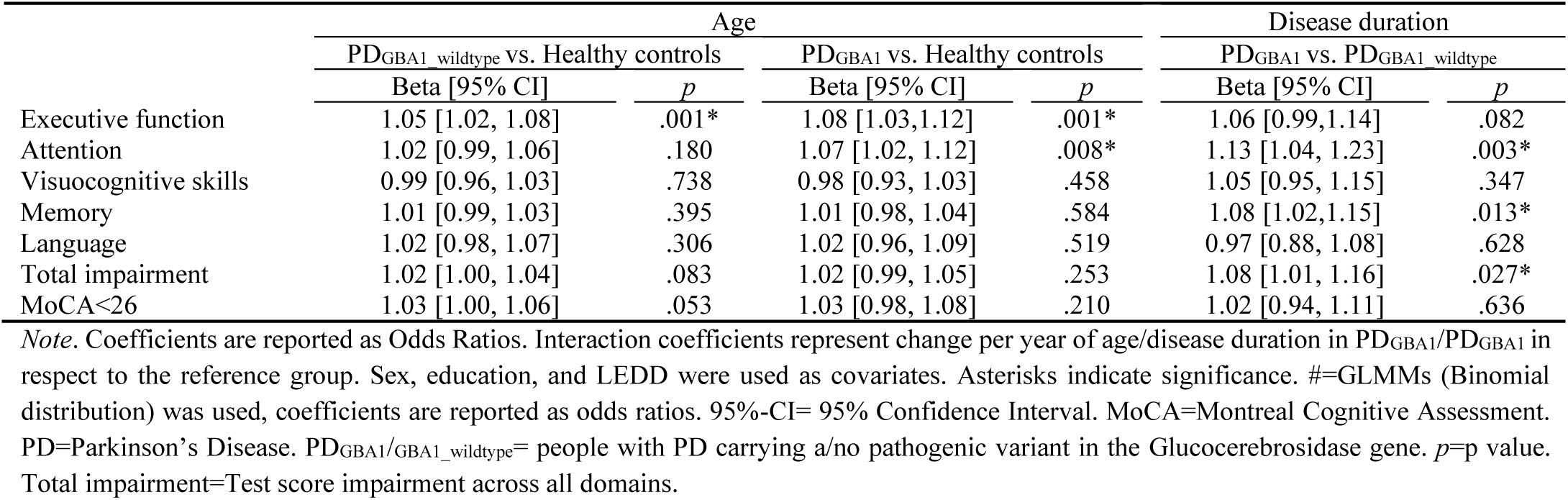
Results of binomial generalized linear mixed effects models (GLMMs) predicting impaired performance (at least one z≤ -1.5) at domain-level and global cognition (MoCA<26) by time*study group interactions across chronological age and disease duration.

### 3.3 Aim 2: Classification of clinical relevance (impaired performance)

Binomial regressions compared the probabilities of demonstrating clinically relevant impaired performance at any available visit. Overall, 54% of PD_GBA1_, 55% of PD_GBA1_wildtype_, and 28% of healthy controls developed impaired performance in the MoCA (Figure 2.c.1, Table S7). Across individual functions, impaired peformance was developed by 15% (working memory) to 55% (processing speed) of the PD_GBA1_ group, by 8% (working memory) to 47% (discrimination) of the PD_GBA1_wildtype_ group, and by 1% (set shifting) to 23% (discrimination) of the healthy controls. Both PD_GBA1_ and PD_GBA1_wildtype_ were more likely to develop impaired performance than healthy controls across all cognitive functions (*p*<.05). PD_GBA1_ were more likely to develop impaired performance than PD_GBA1_wildtype_ across all functions except for the MoCA and visuoconstruction.

Longitudinal trajectories of clinically relevant impaired performance at domain-level (at least one *z*≤-1.5 in cognitive domain), overall cognition-level (*z*≤-1.5 in in any domain), and global cognition (MoCA<26) were compared between study groups using GLMMs (Table S8-S9). Using age as time scale (Figure 2.c.2), both PD_GBA1_ and PD_GBA1_wildtype_ showed a steeper increase in the probability of executive function domain impairment than healthy controls. PD_GBA1_ showed pronounced increase in the probability of attention domain impairment. Across all groups, the probability of memory, visuocognitive skills, and global impairment (MoCA) increased with advancing age. Using disease duration as time scale (Figure 2.c.3), PD_GBA1_ showed a steeper increase in the probability of attention, memory, and overall domain impairment. Across both PD groups, the probability of executive function, visuocognitive skills, and global impairment (MoCA) increased with advancing disease duration.

### 3.4 Aim 3: Associations between different cognitive functions (test performance)

Correlational analyses (Figure 3) between test performance were run within each study group. Moderate within-domain correlations (*r*≥0.30; all *p*<.05) appeared across all groups, involving fluency measures (semantic and phonemic), memory functions (learning, delayed recall, retention, and discrimination), and attentional-executive domains (working memory with phonemic fluency, set shifting with processing speed and working memory). In contrast, moderate cross-domain correlations (*r*≥0.30; all *p*<.05) were observed only within PD groups, where attentional-executive functions correlated with learning, delayed recall, and visuocognitive skills. Additionally, in PD_GBA1_, visuoperception and processing speed correlated with visuoconstruction. Furthermore, moderate correlations of the MoCA (*r*≥0.30; all *p*<.05) were observed with memory (learning, recall) and language among all study groups. However, in PD groups, the MoCA correlated moderately with all other cognitive functions, except fluency measures.

**Figure 3.**
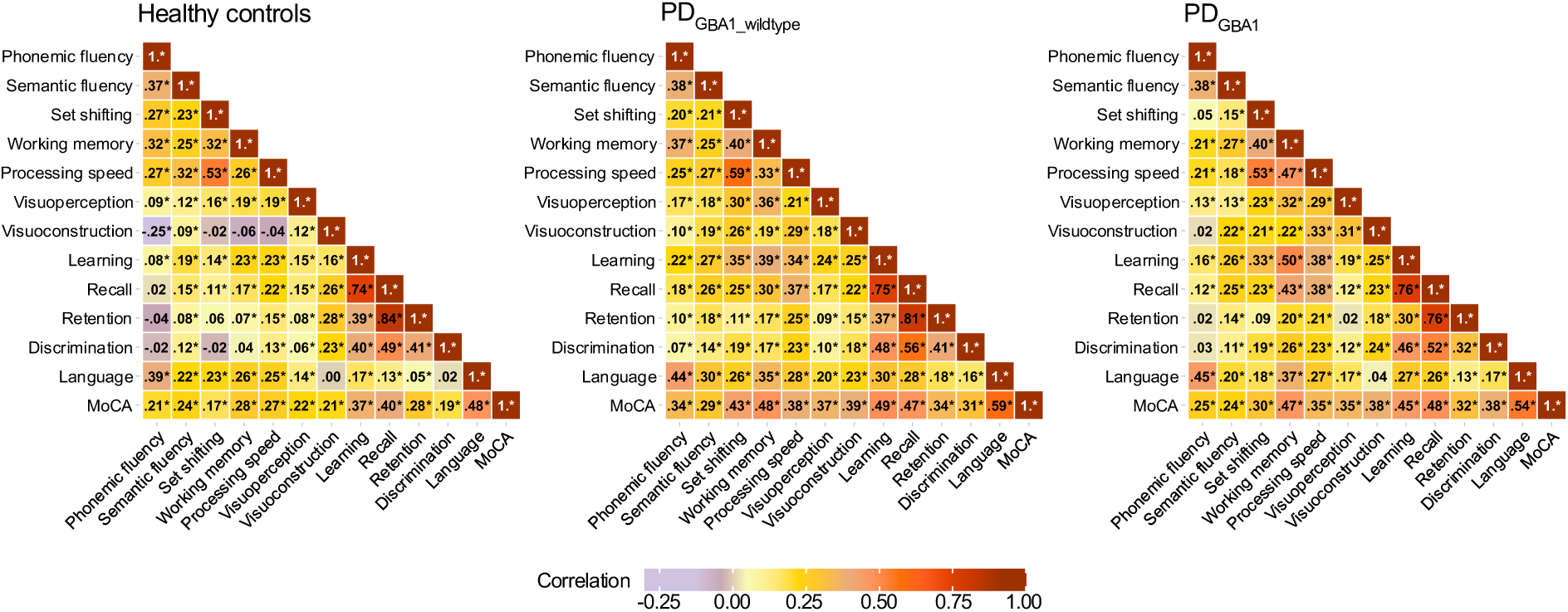
Correlation between cognitive functions (Aim 3) *Note.* The correlograms show the strength of correlations among cognitive functions across all available visits within each study group. Asterisks indicate *p*<.05.

### 3.5 Aim 4: Timing and onset of clinical impairment in PD_GBA1_ vs. PD_GBA1_wildtype_ (impaired performance)

Time-to-first-event models (Table 4, Figure 4), using disease duration as time scale, indicated that PD_GBA1_ showed significantly higher risks of impaired performance across all cognitive functions than PD_GBA1_wildtype_. The highest HRs were observed in working memory, set shifting, and visuoperception. The models showed good to strong discrimination (C=0.72–0.83), except for visuoconstruction, which showed acceptable discrimination (C=0.68).

**Figure 4.**
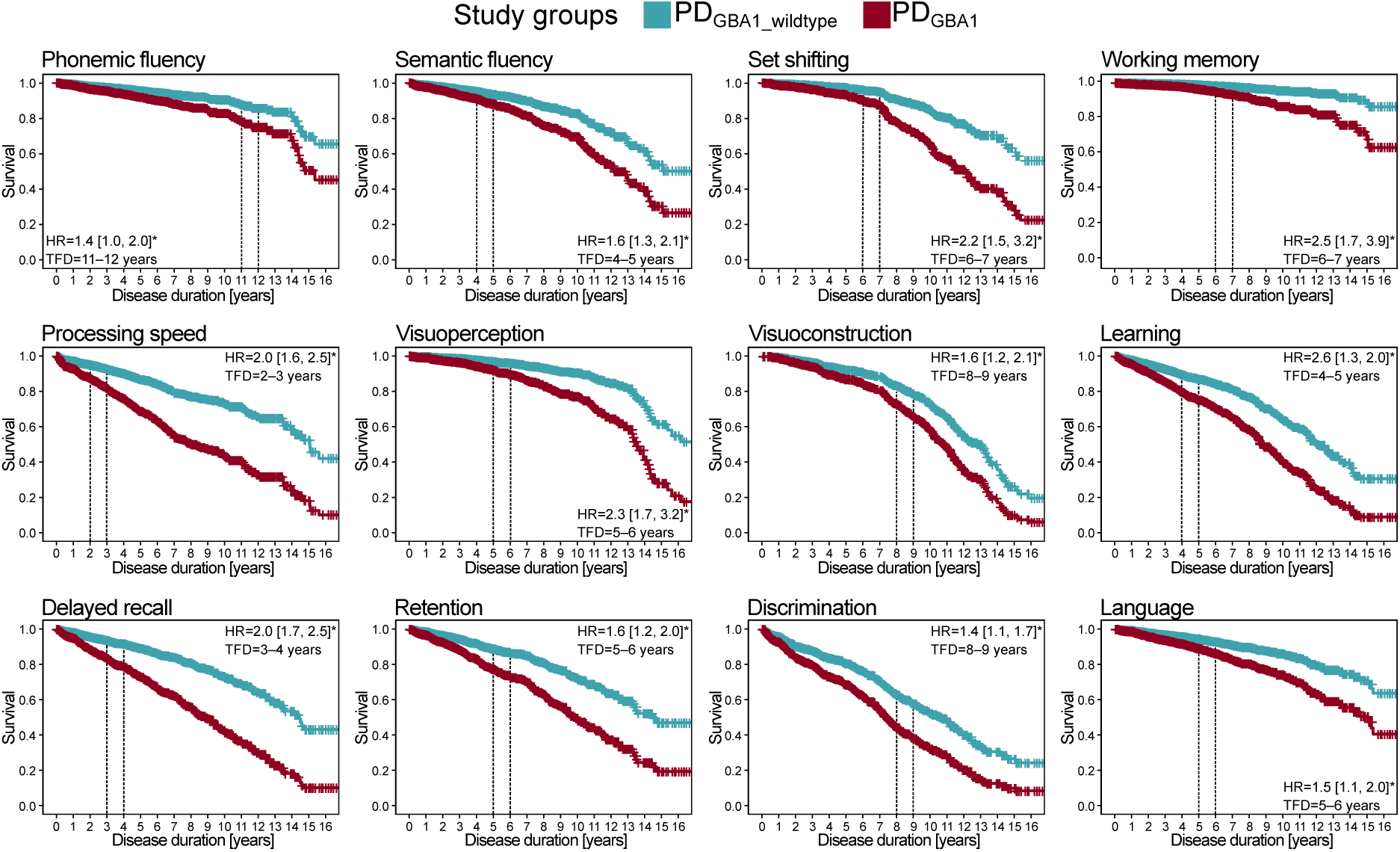
Kaplan-Meier of time-to-first-event models comparing the risk of cognitive test impairment (*z*≤-1.5) in cognitive functions across disease duration between PD_GBA1_ and PD_GBA1_wildtype._ (Aim 4) *Note.* Asterisks indicate *p*<.05. HR=Hazard ratio, with brackets representing 95%-Confidence Intervals. TFD=Time of First Deviation, as indicated by the expanding-time-window analyses.

**Table 4.**
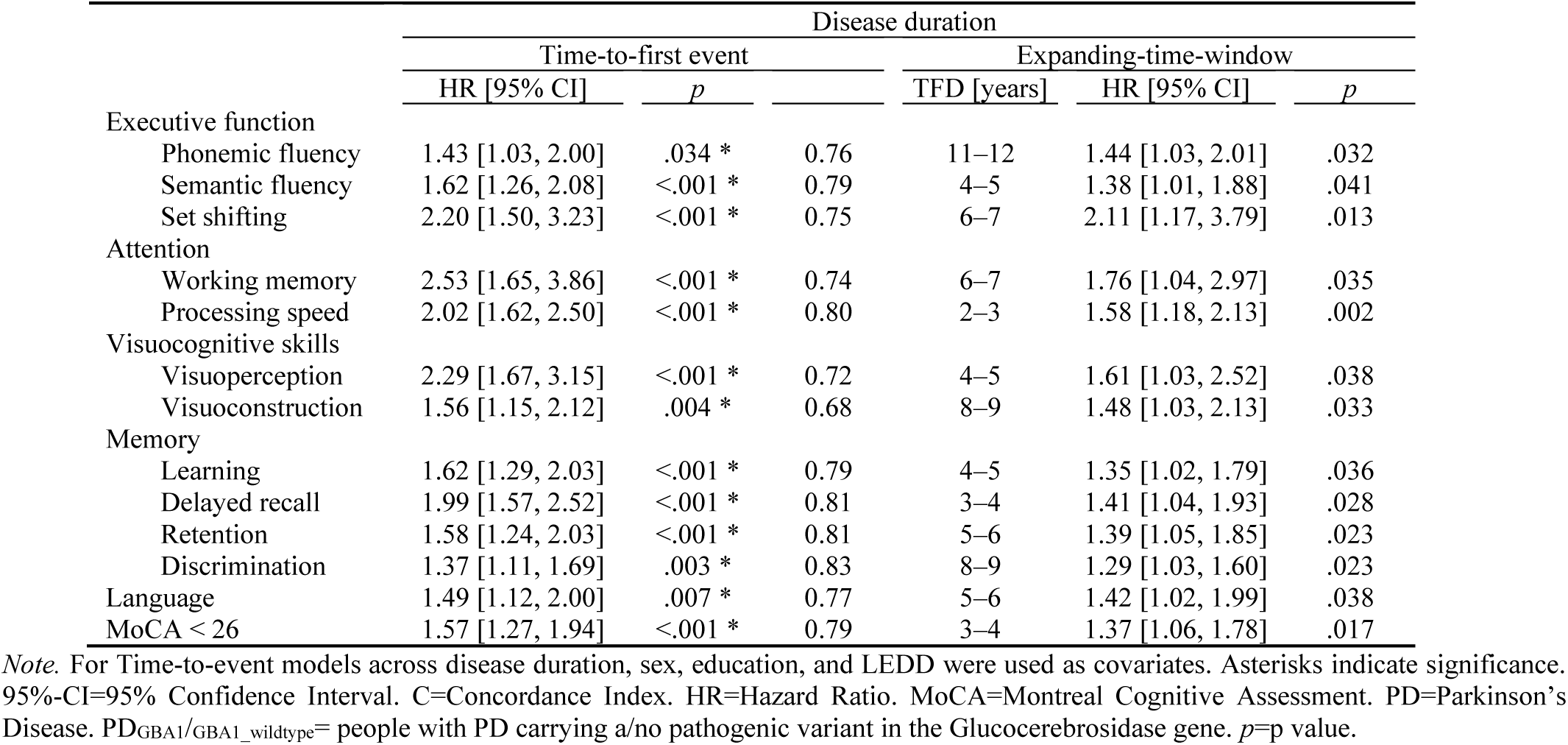
Time-to-first-event models predicting impaired performance (*z≤* -1.5, MoCA<26) in cognitive functions across disease duration.

Next, an expanding time-window approach was applied to each time-to-first-event model, to determine when the risk of impaired performance in PD_GBA1_ first began to discriminate from PD_GBA1_wildtype_ (Table 4, Figure 4). Using 1-year steps across disease duration (0, 1, …, 12 years), the earliest risk elevation in PD_GBA1_ was observed in processing speed (TFD=2–3 years), followed by delayed recall (TFD=3–4 years), global cognition (MoCA<26) semantic fluency, visuoperception, learning (TFD=4-5 years), retention and language (TFD=5–6 years), set shifting, working memory (TFD=6–7 years), visuoconstruction, language (TFD=8–9 years), and phonemic fluency (TFD=11–12 years).

## 4. Discussion

In the present study, we characterized the longitudinal cognitive profile of PD_GBA1_ relative to PD_GBA1_wildtype_ and healthy controls across both disease duration and chronological age. PD_GBA1_ showed widespread cognitive decline across multiple domains (Aim 1), which was not only statistically, but also, importantly, also clinically relevant (Aim 2). Correlational analyses demonstrated interrelationships across cognitive domains, particularly in PD, indicating that cognitive test performance on one test affected performance on other tests (Aim 3). Time-to-first-event models with additional expanding-time-window analysis indicated that the risk of impaired performance was increased across all cognitive functions in PD_GBA1_ beginning 2 to 3 years post-diagnosis (Aim 4).

### Cognitive phenotype in PD_GBA1_

While it is recognized that PD_GBA1_ patients show a more severe decline in global cognition(2,9), knowledge of the progression of individual cognitive functions remains sparse. In this study, we found that clinically relevant impairment in PD_GBA1_ emerged earlier and was more likely to occur across the entire cognitive profile, and that average decline rates were increased in several cognitive domains (attention, visuocognitive skills, memory). These results suggest that PD_GBA1_ represents both an accelerated variant and a partially distinct cognitive phenotype of PD_GBA1_wildtype_(12,22).

In particular, the LMMs showed that cognition in PD_GBA1_ declined more steeply/rapidly than in PD_GBA1_wildtype_, particularly in attention (working memory, processing speed), visuoperception, semantic fluency, and memory (learning, delayed recall, discrimination). Clinically relevant impairment appeared most notably in attention and memory. This cognitive profile closely resembles than seen in Dementia with Lewy Bodies (DLB)(38), indicating early cortical involvement. Presence of pathogenic GBA1 variants ranks among the highest risk factors for Lewy Body Dementia(38), and both conditions demonstrate convergent neuroimaging patterns(39). Interestingly, memory discrimination decline, reported as a less typical feature for Lewy body dementia(40), was found in our expanding-time-window analyses to emerge comparably late in the disease and in proximity to visuoconstruction and language impairment. This timing potentially suggests features of the “posterior-cortical” PDD profile(41). Our results, therefore, indicate that cognitive decline associated with PD_GBA1_ may incorporate features of both DLB and PDD at later stages.

### Time course of cognitive impairment in PD_GBA1_

*Processing speed* declined more significantly more steeply in PD_GBA1_ and was the earliest cognitive function to show elevated impaired performance (2–3 disease years). These results suggest that processing speed impairments may emerge earlier in PD_GBA1_ than previously recognized, a finding reported by only two studies(8,19). Of note, processing speed tests often incorporate motor components (e.g., rapid dot-connection in the TMT-A), which could indirectly reflect dopaminergic integrity(42). Unlike previous analyses(15,20), we controlled for differences in LEDD in our statistical models,. This methodological strength in our study may have uncovered group differences in processing speed that would otherwise have been masked by medication effects on motor function.

PD_GBA1_ declined more steeply in both *memory* capacity and discrimination in our study. Memory capacity impairment (delayed recall: 3–4 disease years, learning: 4–5 disease years, retention: 5–6 disease years) seemed to emerge earlier than discrimination impairment (8–9 disease years). Similarly, prior studies have reported lower memory capacity in PD_GBA1_ vs. PD_GBA1_wildtype_ at ∼6.17 years post-diagnosis(17). Furthermore, longitudinal decline in memory discrimination has been reported beginning ∼8.8 years post-diagnosis(8). When memory capacity alone is impaired, it typically indicates attentional-executive-mediated retrieval deficits, in which patients cannot recall but can recognize presented words(23). Conversely, combined impairment in capacity and discrimination points to encoding deficits (at least partially hippocampally-mediated), in which patients can neither recall nor recognize learned material(23,43,44). Of note, it was reported that PD_GBA1_ commit more encoding errors during memory tasks(45) and exhibit more severe hippocampal dysfunction(13,14,22,39), generally in alignment with our findings. We found moderate-to-strong correlations between attentional-executive functions and memory capacity, but not with discrimination, suggesting greater reliance on other memory functions. In summary, our findings indicate that PD_GBA1_ might first demonstrate retrieval deficits, which later develop into genuine memory impairments(23,43).

Consistent with prior reports(8,15,16), test impairment risk in working memory and set shifting increased at moderate disease duration (6–7 disease years). *Attentional-executive dysfunction* is modulated by dopaminergic denervation in PD and arises from abnormalities in frontal-striatal circuits(46), reported to be more severe in PD_GBA1_ beginning from ∼1.5 years post-diagnosis(22,39). Our result of a later risk elevation of attentional-executive impairment could reflect adaptive nigrostriatal plasticity in PD_GBA1_ before overt clinical manifestation(39). This hypothesis has been supported by studies that have paradoxically found increased dopaminergic binding and altered neural activation patterns even in non-manifesting GBA1 carriers(39). Additionally, given that both working memory and set shifting correlated moderately-to-strongly with processing speed, a core component of cognitive function(23), decreased performance could at least partially result from slowed processing speed.

Visuospatial impairment risk increased during moderate-to-late disease years in PD_GBA1_ (visuoperception: 5–6 disease years, visuoconstruction: 9–10 disease years). These findings align with neuroimaging evidence of more severe posterior-occipital dysfunction in PD_GBA1_ compared with PD_GBA1_wildtype_(39). To the best of our knowledge, visuoconstructive abilities in PD_GBA1_ have been examined in only two studies, with no evidence of performance decrements at ∼2.4 or ∼6.2 years post-diagnosis(12,17). Conversely, both longitudinal studies(8,12) and cross-sectional studies support decreased visuoperception in PD_GBA1_ at ∼2.4 years post-diagnosis (15,18,20), and it has been reported that PD_GBA1_ patients are more prone to develop visual hallucinations(2). We also found that only visuoperception, but not visuoconstruction, declined more steeply in PD_GBA1_ than PD_GBA1_wildtype_, and that visuoperception was the only function in PD_GBA1_ to correlate moderately with visuoconstruction. These findings suggest that visuoperception deficits could dominate visuocognitive skill performance in this population.

For the first time, we observed that *semantic fluency* declined more steeply in PD_GBA1_. This has not been reported in longitudinal studies(8,12) nor in cross-sectional studies before(12,15–18,20). While semantic fluency tasks are sensitive to demographic differences(47), only two cross-sectional studies(17,20)(mean disease duration of ∼3.5 years and ∼6.2 years) of the available studies(8,12,15–18,20), have used normative scores for semantic fluency in PD_GBA1_. In our study, the risk of semantic fluency impairment increased during moderate disease duration and became clinically relevant after 5–6 disease years. Similarly, risk of *language* impairment was increased during later disease years (9–10 disease years) in our study. Prior studies(12–14,48) were limited to earlier disease years (up to ∼6.17 years), possibly explaining the absence of group effects. Our findings underline the need to consider the timing of decline onset across different cognitive functions over disease duration.

Unlike a prior longitudinal study(8), our findings showed neither a steeper decline in phonemic fluency nor a greater probability of clinical impairment. However, according to time-to-first-event analysis, PD_GBA1_ were at greater risk of clinical impairment, diverging relatively late in the disease (11–12 disease years). Given that most visits in our study were conducted at earlier disease durations, it might be possible that the case number was too low at later stages to detect group differences.

We also observed within-group variation of longitudinal trajectories in the models. Impaired performance occurred in up to half of the PD_GBA1_ group. Hence, it appears that GBA1 variant status alone does not seem to determine cognitive prognosis. For instance, a prior analysis of this cohort also indicated that Apolipoprotein e4 status might modulate cognitive progression in PD_GBA1_(49). In this regard, our analyses provide different clinical perspectives on cognitive progression in PD_GBA1_. GLMMs reflect population-level averaged trajectories, including both individuals with severe and benign trajectories, and are thus useful when higher diagnostic accuracy is needed. Binomial regression and time-to-first-event analyses of impaired performance may aid time-critical decisions and prognosis by focusing on the occurrence and onset of clinical milestones.

## Limitations

It might have been possible that people with PD_GBA1_ could have received their PD diagnosis earlier due to their more severe disease progression, implying that group differences might be underestimated. The possibility of selection bias cannot be ruled out, as cognitively more impaired individuals might be more likely to drop-out of studies, as shown in a prior analysis of this cohort(50). While we carefully matched cognitive functions across the cohorts, neuropsychological batteries did not overlap entirely and changed with study protocol adaptations(26,28), resulting in missing values in set shifting and visuoconstruction, and decreased power in these functions as a result. There was a slightly greater proportion of women in the PD_GBA1_ group. We included sex as a covariate, which should have corrected for sex-specific effects on cognitive progression in PD_GBA1_(8).

## Implications

The cognitive phenotype in PD_GBA1_ is generally characterized by more rapid progression across the entire cognitive profile. Early therapeutic interventions and a known GBA1 variant status are time-critical factors for disease modification in PD_GBA1_. Attention, visuoperception, memory, and semantic fluency, are particularly affected in PD_GBA1_ and might be useful in clinical routine and trial monitoring. Prospectively, additional genetic, biological, and sociodemographic factors distinguishing malignant from benign trajectories in PD_GBA1_ may provide new therapeutic opportunities.

## Acknowledgements

PPMI – a public-private partnership – is funded by the Michael J. Fox Foundation for Parkinson’s Research and funding partners, including 4D Pharma, Abbvie, AcureX, Allergan, Amathus Therapeutics, Aligning Science Across Parkinson’s, AskBio, Avid Radiopharmaceuticals, BIAL, BioArctic, Biogen, Biohaven, BioLegend, BlueRock Therapeutics, Bristol-Myers Squibb, Calico Labs, Capsida Biotherapeutics, Celgene, Cerevel Therapeutics, Coave Therapeutics, DaCapo Brainscience, Denali, Edmond J. Safra Foundation, Eli Lilly, Gain Therapeutics, GE HealthCare, Genentech, GSK, Golub Capital, Handl Therapeutics, Insitro, Jazz Pharmaceuticals, Johnson & Johnson Innovative Medicine, Lundbeck, Merck, Meso Scale Discovery, Mission Therapeutics, Neurocrine Biosciences, Neuron23, Neuropore, Pfizer, Piramal, Prevail Therapeutics, Roche, Sanofi, Servier, Sun Pharma Advanced Research Company, Takeda, Teva, UCB, Vanqua Bio, Verily, Voyager Therapeutics, the Weston Family Foundation and Yumanity Therapeutics. Protocol information for The Parkinson’s Progression Markers Initiative (PPMI) Clinical - Establishing a Deeply Phenotyped PD Cohort AM 3.2. can be found on protocols.io or by following this link: https://dx.doi.org/10.17504/protocols.io.n92ldmw6ol5b/v2. This analysis used data from PPMI made available after either a pre-defined embargo or investigator submission of an associated cloud transfer request to the PPMI Data Access Committee.

We would like to thank all participants of the Luxembourg Parkinson’s Study for their important support to our research. Furthermore, we acknowledge the joint effort of the National Centre of Excellence in Research on Parkinson’s Disease (NCER-PD) Consortium members from the partner institutions Luxembourg Centre for Systems Biomedicine, Luxembourg Institute of Health, Centre Hospitalier de Luxembourg, and Laboratoire National de Santé generally contributing to the Luxembourg Parkinson’s Study as listed below: Mariella GRAZIANO 7, Alexandre BISDORFF 5, Rene DONDELINGER 5, Elodie THIRY 3, Gelani ZELIMKHANOV 3, Guy BERCHEM 3, Liliana VILAS BOAS 3, Linda HANSEN 3, Martine GOERGEN 3, Nancy DE BREMAEKER 3, Nico DIEDERICH 3, Romain NATI 3, Roxane BATUTU 3, Sylvia HERBRINK 3, Jochen KLUCKEN 1,3, Rejko KRÜGER 1,2,3, Claire PAULY 2,3, Lukas PAVELKA 2,3, Marijus GIRAITIS 2,3, Maria Fernanda NIÑO URIBE 1,3, Achilleas PEXARAS 2, Alexander HUNDT 2, Alexia MENDIBIDE 2, Ana Festas LOPES 2, Angelo FERRARI 2, Brian DEWITT 2, Carlos GAMIO 2, Estelle HENRY 2, Gaël HAMOT 2, Geeta ACHARYA 2, Hermann THIEN 2, Ilsé RICHARD 2, Johanna TROUET 2, Kate SOKOLOWSKA 2, Katy BEAUMONT 2, Laura GEORGES 2, Lorieza CASTILLO 2, Lucie REMARK 2, Maeva MUNSCH 2, Margaux HENRY 2, Maud THERESINE 2, Olga KOFANOVA 2, Olivia ROLAND 2, Pauline LAMBERT 2, Saïda MTIMET 2, Wim AMMERLANN 2, Anne GRÜNEWALD 1, Armin RAUSCHENBERGER 1,2, Clarissa P. C. GOMES 1, Dheeraj REDDY BOBBILI 1, Ekaterina SOBOLEVA 1,3, Elisa GÓMEZ DE LOPE 1, Enrico GLAAB 1, Evi WOLLSCHEID-LENGELING 1, Francoise MEISCH 1, Giuseppe ARENA 1, Ibrahim BOUSSAAD 1, Iñigo YOLDI BERGUA 1, Jens SCHWAMBORN 1, Kirsten ROOMP 1, Laure PAULY 2, 10, Michael T. HENEKA 1, Michele BASSIS 1, Muhammad ALI 1, Jade JABER 1,3, Patricia MARTINS CONDE 1, Patrick MAY 1, Paul WILMES 1, Piotr GAWRON 1, Rebecca TING JIIN LOO 1, Reinhard SCHNEIDER 1, Ruxandra SOARE 1, Sabine SCHMITZ 1, Sarah NICKELS 1, Sascha HERZINGER 1, Sinthuja PACHCHEK 1, Soumyabrata GHOSH 1, Stefano SAPIENZA 1, Valentin GROUES 1, Venkata SATAGOPAM 1, Isabel SCHWANINGER 1, Liyousew BORGA 1, Sijmen VAN SCHAGEN 1, Gabriel MARTINEZ TIRADO 1, Alan CASTRO MEJIA 1, Messaline FOMO 1, Niloofar KHERADBIN 1, Francesca BOSCHI 1, Francesca TERRANOVA 1, Jochen OHNMACHT 2, Anne-Marie HANFF 2, 10, 11, Carlos VEGA 2, Chouaib MEDIOUNI 2, Deborah MCINTYRE 2, Eduardo ROSALES 2, Fozia NOOR 2, Gessica CONTESOTTO 2, Gloria AGUAYO 2, Guilherme MARQUES 2, Jérôme GRAAS 2, Joëlle FRITZ 2, Magali PERQUIN 2, Manon GANTENBEIN 2, Maura MINELLI 2, Michel VAILLANT 2, Myriam ALEXANDRE 2, Myriam MENSTER 2, Olena TSURKALENKO 2, Raquel SEVERINO 2, Sibylle BÉCHET 3, Tainá M. MARQUES 2, Ulf NEHRBASS 2, Victoria LORENTZ 2, Zied LANDOULSI 2, Sonja JÓNSDÓTTIR 2, David BOUVIER 4, Katrin FRAUENKNECHT 4, Michel MITTELBRONN 1, 2, 4, 10, 12, 13, Roseline LENTZ 6, Jean-Paul NICOLAY 9, Nadine JACOBY 8

1 Luxembourg Centre for Systems Biomedicine, University of Luxembourg, Esch-sur-Alzette, Luxembourg, 2 Luxembourg Institute of Health, Strassen, Luxembourg, 3 Centre Hospitalier de Luxembourg, Strassen, Luxembourg, 4 Laboratoire National de Santé, Dudelange, Luxembourg, 5 Centre Hospitalier Emile Mayrisch, Esch-sur-Alzette, Luxembourg, 6 Parkinson Luxembourg Association, Leudelange, Luxembourg, 7 Association of Physiotherapists in Parkinson’s Disease Europe, Esch-sur-Alzette, Luxembourg, 8 Private practice, Ettelbruck, Luxembourg, 9 Private practice, Luxembourg, Luxembourg, 10 Faculty of Science, Technology and Medicine, University of Luxembourg, Esch-sur-Alzette, Luxembourg, 11 Department of Epidemiology, CAPHRI School for Public Health and Primary Care, Maastricht University Medical Centre+, Maastricht, the Netherlands, 12 Luxembourg Center of Neuropathology, Dudelange, Luxembourg, 13 Department of Life Sciences and Medicine, University of Luxembourg, Esch-sur-Alzette, Luxembourg,

## Author Contributions

MB, WP, RK, ILS, and KB conceptualized and designed the study. MB, CP, SRJ, CS, SB, IW, BR, SL, RK, ILS, and KB acquired and analyzed the data. MB, CP, SRJ, CS, WP, RK, ILS, and KB drafted the manuscript or figures.

## Potential conflicts of interest

|  |  |
| --- | --- |
| Bode, M. | No conflicts of interest in relation to this work |
| Pauly, C. | No conflicts of interest in relation to this work |
| Jónsdóttir, S. R. | No conflicts of interest in relation to this work |
| Schulte, C. | No conflicts of interest in relation to this work |
| Becker, S. | No conflicts of interest in relation to this work |
| Wurster, I. | No conflicts of interest in relation to this work |
| Roeben, B. | No conflicts of interest in relation to this work |
| Lerche, S. | No conflicts of interest in relation to this work |
| Poewe, W. | No conflicts of interest in relation to this work |
| Krüger, R. | No conflicts of interest in relation to this work |
| Liepelt-Scarfone, I. | No conflicts of interest in relation to this work |
| Brockmann, K. | No conflicts of interest in relation to this work |

## Acknowledgements

This work was supported by The Michael J. Fox Foundation for Parkinson’s Research (Michael J. Fox Foundation Write Now Initiative Award).

## Data availability

Data used in the preparation of this article from the Parkinson’s Progression Markers Initiative (PPMI) database (https://www.ppmi-info.org/access-data-specimens/download-data), RRID:SCR_006431 was obtained on 2025-05-22 and 2025-01-29. For up-to-date information on the study, visit https://www.ppmi-info.org. PPMI, a public-private partnership, is funded by the Michael J. Fox Foundation for Parkinson’s Research and funding partners. Visit http://www.ppmi-info.org for more details.

Further data used in the preparation of this manuscript were obtained from the National Centre of Excellence in Research on Parkinson’s Disease (NCER-PD). The National Centre of Excellence in Research on Parkinson’s Disease (NCER-PD) is funded by the Luxembourg National Research Fund (FNR/NCER13/BM/11264123). The NCER-PD database is not publicly available as it is linked to the Luxembourg Parkinson’s study and its internal regulations. The NCER-PD Consortium is willing to share its available data. Its access policy was devised based on the study ethics documents, including the informed consent form approved by the national ethics committee. Requests for access to datasets should be directed to the Data and Sample Access Committee by email at.

The baseline ABC-PD study (study period 2014-2017) was funded by a pre-competitive grant from Janssen Research and Development, a division of Janssen Pharmaceutica N.V. The follow-up study (study period 2018-2020) was funded by a grant from The Michael J. Fox Foundation for Parkinsons Research (Grant Nr. 15227). Neither study sponsor was involved in collection, analysis, and interpretation of data, writing of the report, or decision to submit the paper for publication. Janssen Research and Development was only involved in study design of the baseline visit. The ABC-PD database is not publicly available as it is linked to the internal regulations of the University Hospital of Tuebingen. The PI of the study ILS will consider any requests for access to the data reported in this manuscript (including de-identified participant data and the corresponding data dictionary). Due to restrictions imposed by the Ethics Committee of the Medical Faculty of the University of Tübingen, data access must be in accordance with the approved patient consent procedure to protect patient privacy.

Statistical analysis codes used to perform the analyses in this article are shared on Zenodo (DOI: 10.5281/zenodo.17766974).

## Supplementary Material (Data S1.)

### PPMI STUDY COMMITTEES, CORES, AND COLLABORATORS

#### PPMI EXECUTIVE STEERING COMMITTEE

Kenneth Marek, MD^1^ (Principal Investigator); Tanya Simoni, MD^2^; Andrew Siderowf, MD^3^; Caroline Tanner, MD^4^; Thomas F Tropea, DO^1^; Tatiana Foroud, PhD^5^; Lana Chatline, MD^6^; Brit Mollenhauer, MD^7^; Kalpana Merchant, MD^2^; Douglas Galaskc, MD^8^; Christopher Coffey, PhD^9^; Kathleen Poston, MD^10^; Roseanne Dobkin, PhD^11^; Ethan Brown, MD^4^; Roy Alcalay, MD^12^; Dan Weintraub, MD^3^; Emily Flagg, BA^1^; Kimberly Fabrizio, BA^1^

#### PPMI STEERING COMMITTEE

Susan Gressman, MD^13^; Cornelis Blauwendraat, PhD^14^; Paola Casalin, PhD^15^; Sonya Dumanis, PhD^14^; Raymond James, RN^16^; Karl Kieburtz, MD^17^; Sneha Mantri, MS^18^; Werner Poewe, MD^19^; Michael Schwarzschild, MD^20^; John Seibyl^1^, MD; David Standaert, PhD^21^; Duygu Tosun-Turgut, PhD^4^

#### MICHAEL J. FOX FOUNDATION

Sohini Chowdhury, MA^22^; Jamie Eberling, PhD^22^; Mark Frasier, PhD^22^; Leslie Kirsch, EdD^22^; Katie KopiL, PhD^22^; Maggie Kuhl, BA^22^; Alyssa O’Grady, BA^22^; Todd Sherer, PhD^22^; Tawny Willson, MBS^22^

#### PPMI STUDY CORES

*Project Management Core:* Emily Flagg, BA^1^

*Site Management Core*: Tanya Simuni, MD^2^; Bridget McMahon, BS^1^

*Data Strategy and Technical Operations*: Craig Stanley, PhD^1^; Kim Fabrizio, BA^1^

*Data Management Core:* Dixie Ecklund, MBA^9^, MSN; Christine Kohnen, PhD^9^

*Screening Core:* Tatiana Foroud, PhD^5^; Laura Heathers, BA^5^; Christopher Hobbick, BSCE^5^; Gena Antonopoulos, BSN^5^

*Imaging Core:* John Seibyl, MD^1^; Kathleen Poston, MD^10^

*Statistics Core:* Christopher Coffey, PhD^9^; Chelsea Caspell-Garcia, MS^9^; Michael Brumm, MS^9^

*Bioinformatics Core:* Arthur Toga, PhD^23^; Karen Crawford, MLIS^23^

*Biorepository Core:* Tatiana Foroud, PhD^5^; Jan Hamer, BS^5^

*Biologics Review Committee:* Brit Mollenhauer, MD^7^; Doug Galasko, MD^8^; Kalpana Merchant, MD^2^

*Genetics Core:* Andrew Singleton, PhD^24^

*Pathology Core:* Tatiana Foroud, PhD^5^; Dirk Keene, MD^5^

*Found:* Caroline Tanner, MD^4^; Ethan Brown, MD^4^

*PPMI Online:* Carlie Tanner, MD^4^; Ethan Brown, MD^4^; Lana Chahine, MD^6^; Roseann Dobkin, PhD^11^; Monica Korell, MPH^4^

#### PPMI SITE INVESTIGATORS

Neha Prakash MD^1^; Tanya Simuni, MD^2^; Nabila Dahodwala MD^3^; Caroline Tanner, MD^4^; Lana Chahine MD^6^; Brit Mollanhauer MD^7^; Sebastian Schade MD^7^; Douglas Galasko, MD^8^. Anat Mirelman PhD^12^; Roy Alcalay MD^12^; Katherine Leaver MD^13^; Marie Saint-Hilaire MD^16^; Ruth Schneider MD^17^ Christopher Tarolli MD^17^; Werner Poewe, MD^19^; Aleksandar Videnovic MD^20^; David Standaert PhD^21^; Marissa Dean, MD^21^; Sonja Jonsdottir PhD^25^; Rajko Krueger MD^25^; Claire Pauly PhD^25^; Stewart Factor DO^26^; Penelope Hogarth MD^26^; Robert Hauser MD^28^; Amy Amara PhD^29^; Michelle Fullard MD^29^; Cyrus Zabetian MD^30^; Hubert Fernandez MD^31^; Kathrin Brockmann MD^32^; Isabel WursterPhD^32^; Yen Tei PhD^33^; Paolo Barone PhD^34^; Marina Picillo MD^34^; Stuart Isaacson MD^35^; Alberto Espay MD^36^; Eduardo Tolosa PhD^37^; Javier Ruiz Martinez PhD^38^: Leonidas Stefanis PhD^39^; Kelvin Chou MD^40^; Lorraine Kalia MD^41^; Connie Marras PhD^41^; David Grimes MD^42^; Tiago Mestre PhD^42^; Rajesh Pahwa MD^43^; Mark Lew MD^44^; Holly Shill MD^45^; Shyamal Mehta MD^46^; Giulietta Riboldi MD^47^; Nikolaus McFarland PhD^48^; Ron Postuma MD^49^; Zoltan Mari MD^50^ David Ledingham MD^51^; Nicola Pavese PhD^51^; Michele Hu PhD^52^; Norbert Brueggemann MD^53^,; Christine Klein MD^53^; Bastiaan Bloem PhD^54^: Cristina Simonet PhD^55^, Alastair Noyce PhD^55^: Anetta Janzen PhD^56^; David Pedrosa MD^56^ Wolfgang Oertel PhD^56^; Njideka Okubadejo MD^57^ David Shprecher DO^58^ ArjunTarakad MD^59^; Emile Moukheiber MD^60^

#### PPMI SITE COORDINATORS

Joy Antala^1^; Carla Aranda^2^; Karen Williams^2^; Sophia Melton^2^; Karine Benson^2^; Ashwini Ramachandran^3^; Danielle Potts^3^, Grace LaMoure^3^; Ritikha Vengadesh^3^; Ryan Manzler^3^; Jaime Heller^4^; Primi Ranola^4^; Farah Kausar^4^; Sherri Mosovsky^6^; Diana Willeke^7^; Elizabeth Kalinkara-Gomez^7^; Janelle Rodriguez^8^; Nobuko Kemmotsu^8^; May Eshel^12^; Deborah Raymond^13^; Abigail Desrosiers^16^; Raymond James^16^; Lauren Jackson^17^; Iris Egner^19^; Wesley Schlett^20^; Courtney Blair^21^; Lauren Ruffrage^21^; Berenice Sevilla^26^; Barbara Sommerfeld^26^; Dustin Le^27^; Erica Botting^28^; Gabriella Mazur^28^; Daniele Derlein^29^; Evan Doll^29^; Ying Liu^29^; Ciera Cobb^30^ Olivia Masiewicz^30^; Jennifar Mule^31^; Michael Morsillo^31^; Ella Hilt^32^; Aldazier Jakiran^33^; Dominga Valentino^34^; Lisbeth Pennente^35^; Bobbie Stubbeman^36^; Alicia Garrido^37^; Valeria Ravasi^37^; loana Croitoru^38^ Christos Koros^39^; Nikolas Papagiannakis^39^; Frank Ferrari^40^; Mengyu Zheng^41^; Shawna Reddie^42^ Alicia Alejandra^43^; Andrea Gray^43^; Alejandra Valenzuela^44^; Caitlin Goodman^45^; Sara Dresler^46^; Neil Santos^46^; Fahrial Esha^47^; Kyle Rizer^48^; Nadine Zablith^49^; Liliana Dumitrescu^50^; Debra Galley^51^; Victoria Kate Foster^51^; Jamil Razzaque^52^; Madita Grümmer^54^; Yara Krasowski^54^; Natalie Donkor^55^; Elisabeth Sittig^56^; Oluwadamilola Ojo^57^; Kelly Clark^58^; Rory Mahabir^59^; Kori Ribb^60^; Shamera Willoughby^60^

#### INSTITUTIONS AND AFFILIATIONS

1. Institute for Neurodegenerative Disorders; New Haven, CT, USA
2. Northwestern University; Evanston, IL, USA
3. University of Pennsylvania; Philadelphia, PA, USA
4. University of California, San Francisco; San Francisco, CA, USA
5. Indiana University; Indianapolis, IN, USA
6. University of Pittsburgh; Pittsburgh, PA, USA
7. Paracelsus-Elena Klinik; Kassel, Germany
8. University of California, San Diego, San Diego, CA, USA
9. University of Iowa; Iowa City, IA, USA
10. Stanford University; Stanford, CA. USA
11. Rutgers University; New Brunswick, NJ, USA
12. Tel Aviv Sourasky Medical Center; Tel Aviv, Israel
13. Mount Sinai Beth Israel; New York, NY, USA
14. Coalition for Aligning Science; Chevy Chase, MD, USA
15. BioRep; Milan, Italy
16. Boston University School of Medicine; Boston, MA, USA
17. University of Rochester; Rochester. NY, USA
18. Duke University; Durham, NC, USA
19. University of Innsbruck; Innsbruck, Austria
20. Massachusetts General Hospital; Boston. MA. USA
21. University of Alabama at Birmingham; Birmingham. AL, USA
22. The Michael J. Fox Foundation for Parkinson’s Research; New York. NY. USA
23. Laboratory of Neuroimaging (LONI). USC; Los Angeles. CA. USA
24. National Institute on Aging. NIH; Bethesda. MD. USA
25. University of Luxembourg; Esch-sur-Alzette. Luxembourg
26. Emory University; Atlanta. GA. USA
27. Oregon Health and Science University; Portland. OR, USA
28. University of South Florida; Tampa. FL, USA
29. University of Colorado; Aurora, CO, USA
30. VA Puget Sound Health System; Seattle, WA, USA
31. Cleveland Clinic; Cleveland, OH, USA
32. University of Tubingen; Tubingen, Germany
33. Imperial College of London; London. UK
34. University of Salerno; Salerno. Italy
35. Parkinson’s Disease and Movement Disorders Center; Boca Raton, FL, USA
36. University of Cincinnati; Cincinnati, OH, USA
37. Hospital Clinic of Barcelona; Barcelona, Spain
38. Hospital Universitario Donostia; San Sebastian, Spain
39. University of Athens; Athens, Greece
40. University of Michigan; Ann Arbor, MI, USA
41. Toronto Western Hospital; Toronto, Canada
42. The Ottawa Hospital; Ottawa, Canada
43. University of Kansas Medical Center; Kansas City, KS, USA
44. Keck School of Medicine of the University of Southern California; Los Angeles, CA, USA
45. Barrow Neurological Institute; Phoenix, AZ, USA
46. Mayo Clinic Arizona; Scottsdale, AZ, USA
47. NYU Langone Medical Center; New York, NY, USA
48. University of Florida; Gainesville, FL, USA
49. Montreal Neurological Institute and Hospital/McGill; Montreal, QC, Canada
50. Cleveland Clinic-Las Vegas Lou Ruvo Center for Brain Health; Las Vegas, NV, USA
51. Clinical Ageing Research Unit; Newcastle, UK
52. John Radcliffe Hospital Oxford and Oxford University; Oxford, UK
53. University of Luebeck; Luebeck, Germany
54. Radboud University; Nijmegen, Netherlands
55. Queen Mary University of London; London, UK
56. Philipps-University Marburg; Marburg, Germany
57. University of Lagos; Lagos, Nigeria
58. Banner Sun Health Research Institute; Sun City, AZ, USA
59. Baylor College of Medicine; Houston, TX, USA
60. Johns Hopkins University; Baltimore, MD, USA

#### PPMI STUDY FUNDING PARTNER STATEMENT

PPMI - a public-private partnership - is funded by the Michael J. Fox Foundation for Parkinson’s Research and funding partners, including 4D Pharma, Abbvie, AcureX, Allergan, Amathus Therapeutics, Aligning Science Across Parkinson’s, AskBio, Avid Radiopharmaceuticals, BIAL, BioArctic, Biogen, Biohaven, BioLegend, BlueRock Therapeutics, Bristol-Myers Squibb, Calico Labs, Capsida Biotherapeutics, Celgene, Cerevel Therapeutics, Coave Therapeutics, DaCapo Brainscience, Denali, Edmond J. Satra Foundation, Eli Lilly, Gam Therapeutics, GE Healthcare, Genentech, GSK, Golub Capital, Handl Therapeutics, Insitro, Jazz Pharmaceuticals, Johnson & Johnson Innovative Medicine, Lundbeck, Merck, Meso Scale Discovery, Mission Therapeutics, Neurocrine Biosciences, Neuron23, Neuropore, Pfizer, Piramal, Prevail Therapeutics, Roche, Sanofi, Servier, Sun Pharma Advanced Research Company, Takeda, Teva, UCB, Vanqua Bio, Verily, Voyager Therapeutics, the Weston Family Foundation and Yumanity Therapeutics.

## Supplementary Material (Data S2.)

**Table S1.**
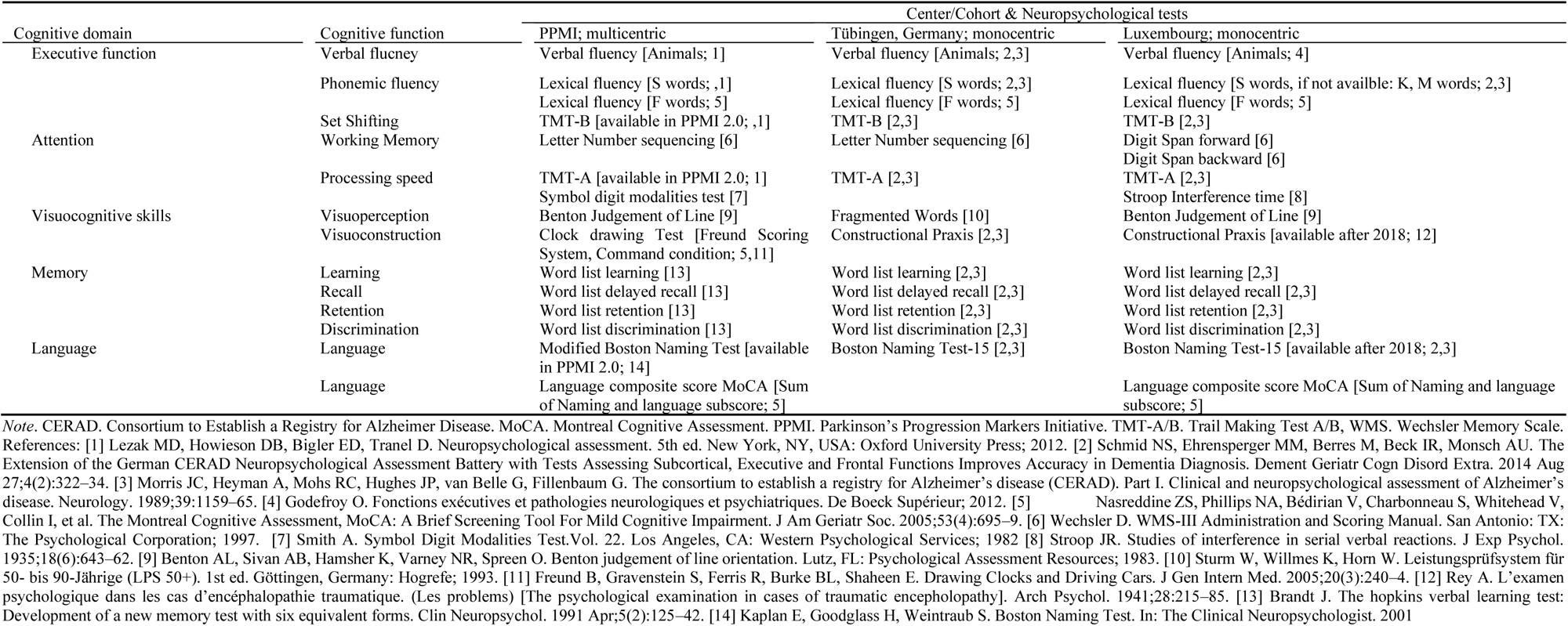
Overview of neuropsychological assessments administered across cohorts mapped to the cognitive function and cognitive domain they assess.

**Table S2.** Published norms used to transfer raw performance of cognitive tests to *z*-scores.

| Cognitive tests | Age |  |
| --- | --- | --- |
|  | Younger | Older |
| <u>Executive function</u> |  |  |
| Animal Fluency (1 min) | Aschenbrenner et al. 2000 (18– 50 years)[1] | Schmid et al. 2014 (50→88 years)[2] |
| Animal Fluency (2 min) | Aschenbrenner et al. 2000 (18– 50 years)[1] |  |
| Lexical fluency (F, S words) | Aschenbrenner et al. 2000 (18– 50 years)[1] | Schmid et al. 2014 (50→88 years)[2] |
| TMT-B | Cavaco et al. (18– 93 years)[3] |  |
| Lexical fluency (K/M words) | Aschenbrenner et al. 2000 (18– 50 years)[1] |  |
| <u>Attention</u> |  |  |
| TMT-A | Cavaco et al. (18– 93 years)[3] |  |
| Letter Digit sequencing [WAIS-III] | Wechsler 1997 (16→89 years)[4] |  |
| Symbol Digit Modalities | Fellows & Schmitter-Edgecombe 2020 (18– >91 years)[5] |  |
| Digit span forward [WMS-III] | Wechsler 1997 (16→89 years)[6] |  |
| Digit span backwards | Wechsler 1997 (16→89 years)[6] |  |
| Stroop Interference Time | van der Elst et al. 2006 (24→81 years)[7] |  |
| <u>Visuocognitive skills</u> |  |  |
| CERAD Construction | Schmid et al. 2014 (50→88 years)[2] |  |
| Benton Judgement of Line Orientation | Calvo et al. 2013 (18–49 years)[8] | Peña-Casanova et al. 2009 (>49→80 years)[9] |
| REY Construction | Fastenau et al. 1999 (19→80 years)[10] |  |
| LPS 50+ | N/A | Sturm et al. 1993 (50→90 years)[11] |
| Clock Drawing | Standardization based on healthy controls from PPMI (<60 years) | Standardization based on healthy controls from PPMI (≥60 years) |
| <u>Memory</u> |  |  |
| CERAD Word list learning total | Hankee et al. (24→78 years)[12] |  |
| CERAD Word list delay | Hankee et al. (24→78 years)[12] |  |
| CERAD retention % | Hankee et al. (24→78 years)[12] |  |
| CERAD discrimination | Hankee et al. (24→78 years)[12] |  |
| HVLT Word list learning total | Benedict et al. 1998 & Duff 2016 (16 →80 years)[13] |  |
| HVLT Word list delay | Benedict et al. 1998 & Duff 2016 (16 →80 years)[13] |  |
| HVLT retention % | Benedict et al. 1998 & Duff 2016 (16 →80 years)[13] |  |
| HVLT discrimination | Benedict et al. 1998 & Duff 2016 (16 →80 years)[13] |  |
| <u>Language</u> |  |  |
| BNT | Tombaugh 1999 (16→95 years)[15] |  |
| Language composite score | Standardization based on healthy controls from PPMI (<60 years) | Standardization based on healthy controls from PPMI (≥60 years) |

**Table S3.**
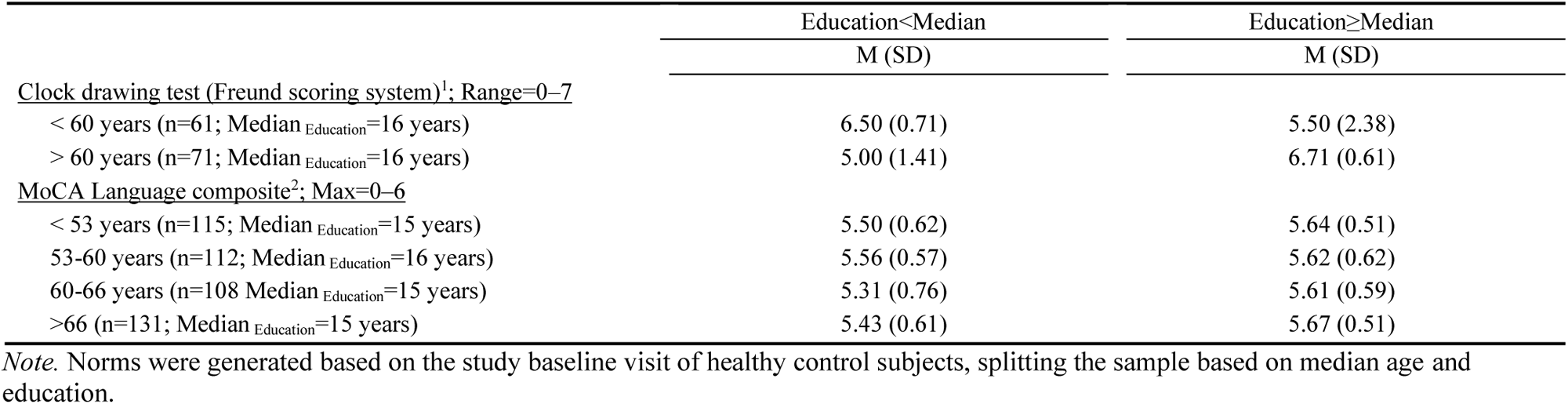
Internal norms used to transfer raw performance of cognitive tests to z-scores.

|  | Education<Median | Education≥Median |
| --- | --- | --- |
|  | M (SD) | M (SD) |
| <u>Clock drawing test (Freund scoring system)<sup>1</sup>; Range=0–7</u> |  |  |
| < 60 years (n=61; Median<sub>Education</sub>=16 years) | 6.50 (0.71) | 5.50 (2.38) |
| > 60 years (n=71; Median<sub>Education</sub>=16 years) | 5.00 (1.41) | 6.71 (0.61) |
| <u>MoCA Language composite<sup>2</sup>; Max=0–6</u> |  |  |
| < 53 years (n=115; Median<sub>Education</sub>=15 years) | 5.50 (0.62) | 5.64 (0.51) |
| 53–60 years (n=112; Median<sub>Education</sub>=16 years) | 5.56 (0.57) | 5.62 (0.62) |
| 60–66 years (n=108 Median<sub>Education</sub>=15 years) | 5.31 (0.76) | 5.61 (0.59) |
| >66 (n=131; Median<sub>Education</sub>=15 years) | 5.43 (0.61) | 5.67 (0.51) |

*Note.* Norms were generated based on the study baseline visit of healthy control subjects, splitting the sample based on median age and education.

**Figure S1.**
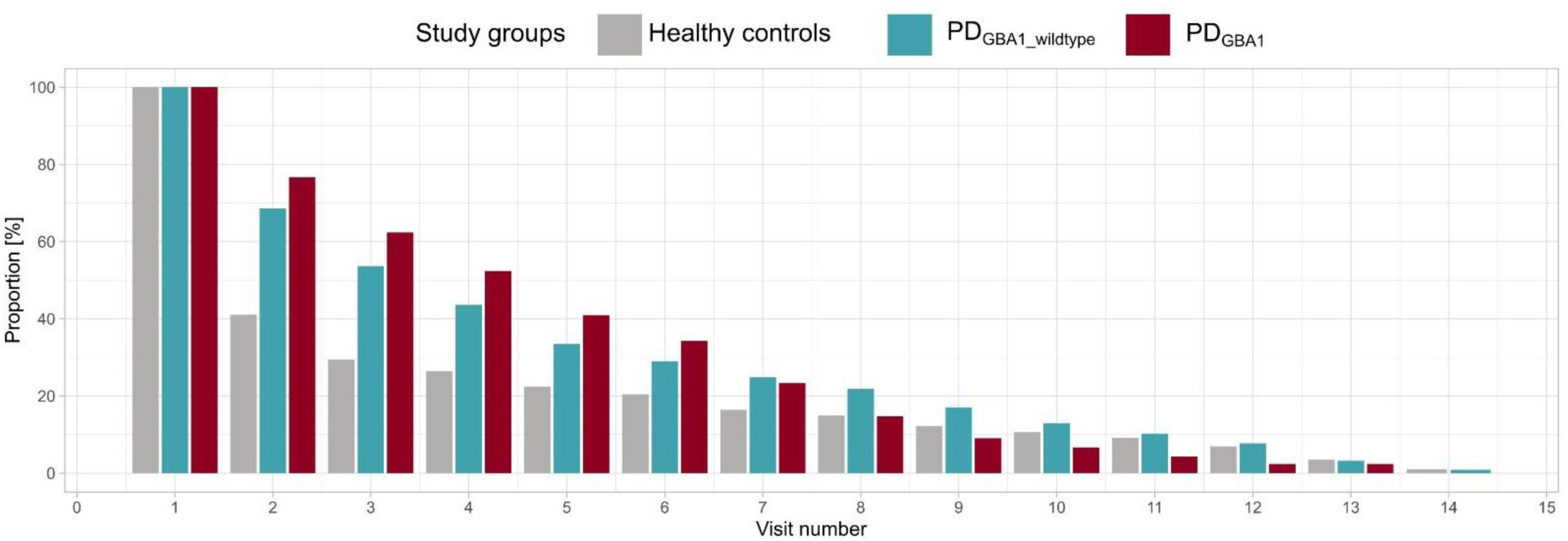
**Longitudinal retention of study participants across (follow-up) visits** *Note.* Bars represent percentage of original sample (*n* = 548 healthy controls, *n* = 906 PD_GBA1_wildtype_, *n* = 210 PD_GBA1_) remaining at each assessment time point. Visit 1 represents baseline visit, with subsequent visits representing follow-up assessments.

**Table S4.** Results of Linear mixed effects models (LMMs) predicting cognitive performance (*z-*score, MoCA score) across chronological age for PD_GBA1_ and PD_GBA1_wildtype_ to healthy controls.

|  | Intercept |  | Age |  | PD<sub>GBA1_wildtype</sub> |  | PD<sub>GBA1</sub> |  | Age* PD<sub>GBA1_wildtype</sub> |  | Age*PD<sub>GBA1</sub> |  |
| --- | --- | --- | --- | --- | --- | --- | --- | --- | --- | --- | --- | --- |
|  | Beta [95% CI] | p | Beta [95% CI] | p | Beta [95% CI] | p | Beta [95% CI] | p | Beta [95% CI] | p | Beta [95% CI] | p |
| Executive function |  |  |  |  |  |  |  |  |  |  |  |  |
| Phonemic fluency | -1.51 [-2.05,-0.96] | <.001 * | 0.03 [ 0.02, 0.03] | <.001 * | 0.18 [-0.32, 0.68] | .474 | 0.75 [-0.04, 1.55] | .065 | -0.01 [-0.01, 0.00] | .065 | -0.02 [-0.03, 0.00] | .010 * |
| Semantic fluency | -0.59 [-1.02,-0.16] | .008 * | 0.01 [ 0.00, 0.01] | .016 * | 0.60 [ 0.12, 1.08] | .015 * | 1.55 [ 0.78, 2.31] | <.001 * | -0.01 [-0.02, 0.00] | .002 * | -0.03 [-0.04,-0.02] | <.001 * |
| Set shifting | -0.41 [-1.08, 0.26] | .228 | 0.01 [ 0.00, 0.02] | .008 * | 2.13 [ 1.48, 2.78] | <.001 * | 0.27 [-0.96, 1.50] | .672 | -0.04 [-0.05,-0.03] | <.001 * | -0.01 [-0.03, 0.00] | .148 |
| Attention |  |  |  |  |  |  |  |  |  |  |  |  |
| Working memory | -1.10 [-1.51,-0.68] | <.001 * | 0.01 [ 0.00, 0.01] | .002 * | 0.74 [ 0.31, 1.17] | .001 * | 1.52 [ 0.84, 2.21] | <.001 * | -0.01 [-0.02,-0.01] | <.001 * | -0.03 [-0.04,-0.02] | <.001 * |
| Processing speed | -2.39 [-2.95,-1.83] | <.001 * | 0.04 [ 0.03, 0.04] | <.001 * | 0.59 [ 0.04, 1.14] | .037 * | 1.17 [ 0.29, 2.05] | .010 * | -0.02 [-0.03,-0.01] | <.001 * | -0.04 [-0.05,-0.02] | <.001 * |
| Visuocognitive skills |  |  |  |  |  |  |  |  |  |  |  |  |
| Visuoperception | 0.34 [-0.35, 1.03] | .334 | 0.00 [-0.01, 0.00] | .262 | 0.02 [-0.55, 0.59] | .950 | 0.05 [-0.86, 0.95] | .921 | 0.00 [-0.01, 0.01] | .414 | -0.01 [-0.02, 0.01] | .254 |
| Visuoconstruction | 3.08 [ 1.74, 4.43] | <.001 * | -0.03 [-0.04,-0.01] | .001 * | 1.55 [ 0.31, 2.80] | .015 * | 0.69 [-1.28, 2.66] | .494 | -0.03 [-0.04,-0.01] | .005 * | -0.02 [-0.05, 0.01] | .230 |
| Memory |  |  |  |  |  |  |  |  |  |  |  |  |
| Learning | -0.42 [-0.95, 0.11] | .121 | 0.00 [-0.01, 0.00] | .242 | 1.06 [ 0.50, 1.61] | <.001 * | 0.84 [-0.04, 1.72] | .062 | -0.02 [-0.03,-0.01] | <.001 * | -0.02 [-0.04,-0.01] | .001 * |
| Delayed recall | 0.09 [-0.58, 0.75] | .796 | -0.01 [-0.01, 0.00] | .058 | 0.58 [ 0.03, 1.13] | .040 * | 0.81 [-0.06, 1.69] | .069 | -0.02 [-0.02,-0.01] | <.001 * | -0.02 [-0.04,-0.01] | .001 * |
| Retention | 0.51 [-0.16, 1.18] | .134 | -0.01 [-0.02, 0.00] | .008 * | 0.24 [-0.30, 0.78] | .379 | 0.39 [-0.46, 1.23] | .370 | -0.01 [-0.02, 0.00] | .042 * | -0.01 [-0.02, 0.00] | .095 |
| Discrimination | -0.07 [-0.69, 0.55] | .825 | 0.00 [-0.01, 0.00] | .222 | 0.32 [-0.26, 0.89] | .282 | 0.69 [-0.20, 1.58] | .127 | -0.01 [-0.02, 0.00] | .016 * | -0.02 [-0.03, 0.00] | .011 * |
| Language | -0.68 [-1.27,-0.09] | .024 * | 0.00 [-0.01, 0.01] | .896 | 0.18 [-0.39, 0.75] | .541 | 0.62 [-0.28, 1.52] | .178 | -0.01 [-0.02, 0.00] | .095 | -0.02 [-0.03, 0.00] | .029 * |
| MoCA score | 27.24 [26.06,28.42] | <.001 * | -0.03 [-0.05,-0.02] | <.001 * | 2.57 [ 1.21, 3.94] | <.001 * | 3.48 [ 1.30, 5.65] | .002 * | -0.06 [-0.08,-0.04] | <.001 * | -0.08 [-0.11,-0.04] | <.001 * |

*Note.* Sex, education, and LEDD were used as covariates. Asterisks indicate significance. 95%-CI= 95% Confidence Interval. MoCA=Montreal Cognitive Assessment. PD=Parkinson's Disease. PD<sub>GBA1</sub>/GBA1\_wildtype= people with PD carrying a/no pathogenic variant in the Glucocerebrosidase gene. *p*=*p* value.

**Table S5.**
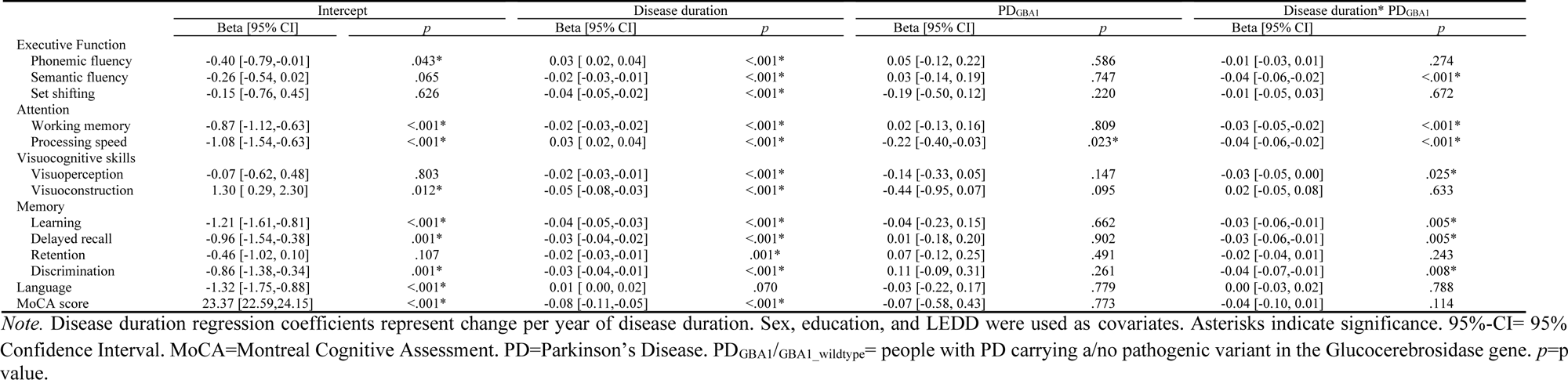
Results of Linear mixed effects models (LMMs) predicting cognitive performance (*z-*score, MoCA score) across disease duration for PD_GBA1_ compared to PD_GBA1_wildtype_. *Note.* Disease duration regression coefficients represent change per year of disease duration. Sex, education, and LEDD were used as covariates. Asterisks indicate significance. 95%-CI= 95%

**Table S6.** Intraclass correlation [ICC] of Linear mixed effects models (LMMs) predicting cognitive performance (*z-*score, MoCA score) across chronological age and disease duration for study groups.

|  | Age |  |  | Disease duration |  |
| --- | --- | --- | --- | --- | --- |
|  | Healthy control | PD<sub>GBA1</sub>_wildtype | PD<sub>GBA1</sub> | PD<sub>GBA1</sub>_wildtype | PD<sub>GBA1</sub> |
| Executive Function |  |  |  |  |  |
| Phonemic fluency | 0.74 | 0.69 | 0.66 | 0.70 | 0.66 |
| Semantic fluency | 0.62 | 0.64 | 0.62 | 0.64 | 0.61 |
| Set shifting | 0.64 | 0.71 | 0.72 | 0.72 | 0.71 |
| Attention |  |  |  |  |  |
| Working memory | 0.74 | 0.71 | 0.65 | 0.71 | 0.66 |
| Processing speed | 0.65 | 0.71 | 0.71 | 0.71 | 0.71 |
| Visuocognitive skills |  |  |  |  |  |
| Visuoperception | 0.51 | 0.63 | 0.64 | 0.63 | 0.63 |
| Visuoconstruction | 0.57 | 0.52 | 0.58 | 0.54 | 0.59 |
| Memory |  |  |  |  |  |
| Learning | 0.63 | 0.62 | 0.55 | 0.63 | 0.60 |
| Delayed recall | 0.66 | 0.65 | 0.64 | 0.66 | 0.65 |
| Retention | 0.54 | 0.51 | 0.43 | 0.51 | 0.44 |
| Discrimination | 0.42 | 0.46 | 0.43 | 0.47 | 0.46 |
| Language | 0.54 | 0.54 | 0.57 | 0.55 | 0.56 |
| MoCA score | 0.43 | 0.40 | 0.39 | 0.82 | 0.83 |

*Note.* The ICC (percentage scale, 0-100%) indicates the proportion of total variance that occurs between clusters/groups, with higher ICC values indicating that observations within the same group are more similar to each other. ICC=Intraclass correlation. MoCA=Montreal Cognitive Assessment. PD=Parkinson's Disease. PD<sub>GBA1</sub>/GBA1\_wildtype= people with PD carrying a/no pathogenic variant in the Glucocerebrosidase gene.

**Table S7.** Results of binomial regressions comparing proportions of occurrence of impaired performance (*z≤* -1.5, MoCA<26) across the whole study period between study groups.

|  | Study group |  |  | Effect size |  |  |
| --- | --- | --- | --- | --- | --- | --- |
|  | Healthy control | PD<sub>GBA1_wildtype</sub> | PD<sub>GBA1</sub> | Healthy control vs. PD<sub>GBA1_wildtype</sub> | Healthy control vs. PD<sub>GBA1</sub> | PD<sub>GBA1_wildtype</sub> vs. PD<sub>GBA1</sub> |
| Executive Function |  |  |  |  |  |  |
| Phonemic fluency | 13% | 19% | 22% | 3.74* | 3.67* | 1.64* |
| Semantic fluency | 11% | 29% | 42% | 8.07* | 12.62* | 2.17* |
| Set shifting | 1% | 17% | 25% | 21.76* | 63.96* | 1.74* |
| Attention |  |  |  |  |  |  |
| Working memory | 2% | 8% | 15% | 8.24* | 11.01* | 2.48* |
| Processing speed | 13% | 36% | 55% | 8.16* | 17.00* | 3.30* |
| Visuocognitive skills |  |  |  |  |  |  |
| Visuoperception | 4% | 16% | 26% | 6.61* | 13.40* | 2.05* |
| Visuoconstruction | 14% | 35% | 40% | 4.05* | 7.44* | 1.27 |
| Memory |  |  |  |  |  |  |
| Learning | 14% | 41% | 48% | 8.14* | 7.28* | 1.68* |
| Delayed recall | 14% | 35% | 45% | 8.81* | 9.01* | 2.32* |
| Retention | 15% | 33% | 40% | 8.23* | 7.28* | 2.11* |
| Discrimination | 23% | 47% | 54% | 7.81* | 5.53* | 1.80* |
| Language | 13% | 25% | 28% | 4.04* | 4.52* | 1.55* |
| MoCA<26 | 28% | 55% | 54% | 5.60* | 6.00* | 1.26 |

*Note.* For comparisons with the healthy control group, sex, education, LEDD, and depression severity were used as covariates. Exponentiated beta coefficients are reported as effect size. Asterisks indicate significance. Coefficients are reported as odds ratios. 95%-CI= 95% Confidence Interval. MoCA=Montreal Cognitive Assessment. PD=Parkinson's Disease. PD<sub>GBA1</sub>/GBA1\_wildtype= people with PD carrying a/no pathogenic variant in the Glucocerebrosidase gene.

**Table S8.** Results of Generalized linear mixed effects models (GLMMs) predicting occurrence of impaired performance (at least one *z≤* -1.5) within each cognitive domain and all domains as well as global cognition (MoCA<26) across chronological age for PD_GBA1_ and PD_GBA1_wildtype_ to healthy controls.

|  | Intercept |  | Age |  | PD<sub>GBA1_wildtype</sub> |  | PD<sub>GBA1</sub> |  | Age* PD<sub>GBA1_wildtype</sub> |  | Age*PD<sub>GBA1</sub> |  |
| --- | --- | --- | --- | --- | --- | --- | --- | --- | --- | --- | --- | --- |
|  | Beta [95% CI] | *p* | Beta [95% CI] | *p* | Beta [95% CI] | *p* | Beta [95% CI] | *p* | Beta [95% CI] | *p* | Beta [95% CI] | *p* |
| Executive function | 0.01 [0.00,0.06] | <.001* | 1.00 [0.98,1.03] | .842 | 0.13 [0.02,0.79] | .027* | 0.04 [0.00,0.59] | .019* | 1.05 [1.02,1.08] | .001* | 1.08 [1.03,1.12] | .001* |
| Attention | 0.52 [0.06, 4.13] | .533 | 0.96 [0.93, 0.99] | .008* | 1.57 [0.17,14.81] | .692 | 0.31 [0.01, 7.72] | .477 | 1.02 [0.99, 1.06] | .180 | 1.07 [1.02, 1.12] | .008* |
| Visuocognitive skills | 0.00 [0.00, 0.00] | <.001* | 1.08 [1.05, 1.12] | <.001* | 1.43 [0.11, 18.97] | .785 | 5.55 [0.19,158.65] | .317 | 0.99 [0.96, 1.03] | .738 | 0.98 [0.93, 1.03] | .458 |
| Memory | 0.06 [0.01, 0.32] | .001* | 1.03 [1.01, 1.05] | .002* | 1.28 [0.30, 5.50] | .743 | 1.87 [0.22,15.61] | .562 | 1.01 [0.99, 1.03] | .395 | 1.01 [0.98, 1.04] | .584 |
| Language<sup>#</sup> | 0.07 [0.01, 0.74] | .027* | 1.02 [0.98, 1.05] | .320 | 0.42 [0.03, 6.44] | .537 | 0.62 [0.01,40.19] | .824 | 1.02 [0.98, 1.07] | .306 | 1.02 [0.96, 1.09] | .519 |
| Total impairment | 0.36 [0.10, 1.28] | .116 | 1.01 [1.00, 1.03] | .101 | 0.98 [0.26, 3.66] | .981 | 1.71 [0.21,13.63] | .612 | 1.02 [1.00, 1.04] | .083 | 1.02 [0.99, 1.05] | .253 |
| MoCA<26 | 0.01 [0.00, 0.04] | <.001* | 1.08 [1.05, 1.11] | <.001* | 0.47 [0.06, 3.65] | .471 | 0.64 [0.03,13.28] | .773 | 1.03 [1.00, 1.06] | .053 | 1.03 [0.98, 1.08] | .210 |

*Note.* Coefficients are reported as odds ratios. Age regression coefficients represent change per year of age. Sex, education, and LEDD were used as covariates. Asterisks indicate significance. <sup>#</sup>=GLMMs (Binomial distribution) was used, coefficients are reported as odds ratios. For reasons of convergence, depression severity was categorized in “not depressed” vs. “depressed”. Asterisks 95%-CI= 95% Confidence Interval. MoCA=Montreal Cognitive Assessment. PD=Parkinson’s Disease. PD<sub>GBA1</sub>/GBA1\_wildtype= people with PD carrying a/no pathogenic variant in the Glucocerebrosidase gene. *p*=*p* value. Total impairment=Test score impairment across all domains.

**Table S9.**
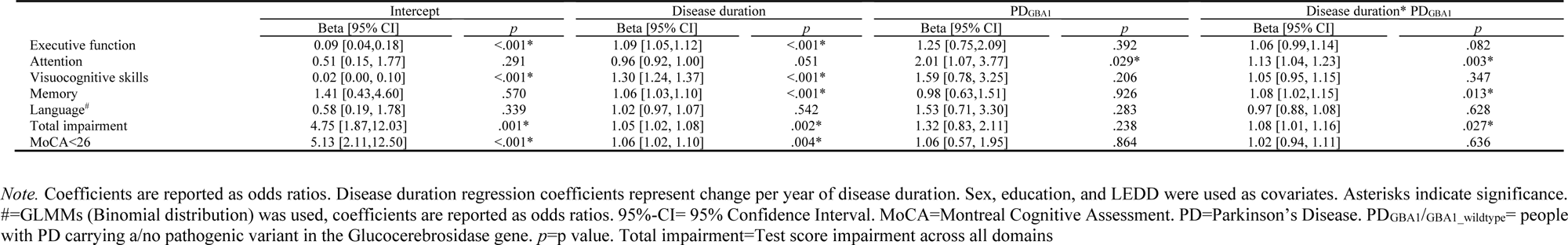
Results of Linear mixed effects models (GLMMs) predicting impaired performance (at least one *z≤* -1.5) within each cognitive domain and all domains as well as global cognition (MoCA<26) for PD_GBA1_.

|  | Intercept |  | Disease duration |  | PD<sub>GBA1</sub> |  | Disease duration* PD<sub>GBA1</sub> |  |
| --- | --- | --- | --- | --- | --- | --- | --- | --- |
|  | Beta [95% CI] | *p* | Beta [95% CI] | *p* | Beta [95% CI] | *p* | Beta [95% CI] | *p* |
| Executive function | 0.09 [0.04,0.18] | <.001* | 1.09 [1.05,1.12] | <.001* | 1.25 [0.75,2.09] | .392 | 1.06 [0.99,1.14] | .082 |
| Attention | 0.51 [0.15, 1.77] | .291 | 0.96 [0.92, 1.00] | .051 | 2.01 [1.07, 3.77] | .029* | 1.13 [1.04, 1.23] | .003* |
| Visuocognitive skills | 0.02 [0.00, 0.10] | <.001* | 1.30 [1.24, 1.37] | <.001* | 1.59 [0.78, 3.25] | .206 | 1.05 [0.95, 1.15] | .347 |
| Memory | 1.41 [0.43,4.60] | .570 | 1.06 [1.03,1.10] | <.001* | 0.98 [0.63,1.51] | .926 | 1.08 [1.02,1.15] | .013* |
| Language<sup>#</sup> | 0.58 [0.19, 1.78] | .339 | 1.02 [0.97, 1.07] | .542 | 1.53 [0.71, 3.30] | .283 | 0.97 [0.88, 1.08] | .628 |
| Total impairment | 4.75 [1.87,12.03] | .001* | 1.05 [1.02, 1.08] | .002* | 1.32 [0.83, 2.11] | .238 | 1.08 [1.01, 1.16] | .027* |
| MoCA<26 | 5.13 [2.11,12.50] | <.001* | 1.06 [1.02, 1.10] | .004* | 1.06 [0.57, 1.95] | .864 | 1.02 [0.94, 1.11] | .636 |

*Note.* Coefficients are reported as odds ratios. Disease duration regression coefficients represent change per year of disease duration. Sex, education, and LEDD were used as covariates. Asterisks indicate significance. <sup>#</sup>=GLMMs (Binomial distribution) was used, coefficients are reported as odds ratios. 95%-CI= 95% Confidence Interval. MoCA=Montreal Cognitive Assessment. PD=Parkinson’s Disease. PD<sub>GBA1</sub>/GBA1\_wildtype= people with PD carrying a/no pathogenic variant in the Glucocerebrosidase gene. *p*=*p* value. Total impairment=Test score impairment across all domains

